# The landscape of functional brain network impairments in late-onset GM2 gangliosidosis

**DOI:** 10.1101/2022.09.11.22279835

**Authors:** D Rangaprakash, Olivia E Rowe, Christopher D Stephen, Florian S Eichler, Eva-Maria Ratai, Robert L Barry

## Abstract

Late-onset GM2 gangliosidosis (LOGG) is an ultra-rare neurological disease with motor, cognitive and psychiatric manifestations. It is caused by mutations in the *HEXA* or *HEXB* genes. Although cerebellar structural and metabolic impairments have been established, global brain functional impairments in this disease remain unknown. In this first functional MRI (fMRI) report on LOGG (N=14), we took an exploratory, multi-pronged approach by assessing impairments in several resting-state fMRI signal characteristics: fMRI signal strength, neurovascular coupling, static and time-varying functional connectivity, and network topology. Contrary to the predominance of cerebellar aberrations in prior non-functional studies, we found more widespread cortical aberrations (*p*<0.05, FDR-corrected) mainly in cognitive control networks but also in the default mode and somatomotor networks. There was reduced fMRI signal strength, enhanced neurovascular coupling, pathological hyper-connectivity, and altered temporal variability of connectivity in the LOGG cohort. We also observed an imbalance between functional segregation and integration as seen in other psychiatric/neurological disorders, with heightened segregation and suppressed integration (i.e., inefficient brain-wide communication). Some of these imaging markers were significantly associated with clinical measures, as well as with *HEXA* and *HEXB* gene expression. These aberrations might contribute to psychiatric symptoms (psychosis, mood disturbances), cognitive impairments (memory, attention, executive function), and oculomotor disturbances commonly seen in LOGG. Future LOGG imaging studies should probe brain function in addition to structure/metabolism while looking for mechanistic insights beyond the cerebellum.

## 1. Introduction

Functional neuroimaging has provided insight into the functional neuroarchitecture of neurological diseases such as Alzheimer’s disease [1], Parkinson’s disease [2] and multiple sclerosis [3]. Blood oxygenation level dependent (BOLD) functional MRI (fMRI) measures the hemodynamic response to energy demands from local neural activity with high spatial resolution and specificity [4]. The last two decades have leveraged this technology to study various aspects of brain activity and connectivity while at rest [5]. Resting-state fMRI, provides us with various dimensions of information, such as co-activation between pairs of brain regions (functional connectivity) [5], topology of whole-brain connectivity networks (complex network models) [6], strength of low-frequency BOLD fluctuations [7], and neurovascular coupling [8].

Despite the rich battery of information available through fMRI, these methods have not been leveraged to probe the mechanistic understanding of late-onset GM2-gangliosidosis (LOGG). LOGG is an ultra-rare form of GM2-gangliosidosis [9] with onset in late teens or adulthood. (It should be noted that LOGG’s clinical course differs considerably from classic infantile-onset [10].) While GM2-related disorders carry a prevalence rate of 1 in 3500, LOGG is 1 in ∼300,000. LOGG occurs mainly, although not exclusively, in persons of Ashkenazi Jewish ancestry (about 10 million global population and 6 million in the US) [11]. LOGG encompasses two closely related but distinguishable diseases, late-onset Tay-Sachs disease (LOTS) and late-onset Sandhoff disease (LOSD) [12]. Neurological dysfunction in LOGG is caused by lysosomal storage dysfunction related to mutations in the α- and β-subunits of β-hexosaminidase (*HEXA* or *HEXB*) [13], and has no treatment or cure. LOGG is mainly characterized by limb weakness and cerebellar ataxia, but cognitive impairments and psychiatric symptoms, including psychosis and mood disorder, have also been reported [14]. Severe psychiatric presentations have been recorded in 40% of LOGG patients [13]. These patients, whose severe symptoms often mimic different psychiatric disorders [15], respond poorly to typical psychiatric drugs [15], which could even worsen/accelerate their neurological symptoms [16]. Cognitive impairment has been mainly reported in LOTS, where it has been observed in up to 44% of cases [14], particularly involving memory and executive functioning [17]. Barritt et al. [18] reported deficits in processing speed, attention, and memory, while another study revealed severe cognitive impairment in 7 of 10 LOGG patients [19]. Another study found that among 63 reported cases, 30–50% exhibited psychosis, >25% exhibited mood disorder, and >20% showed cognitive impairment [20], concluding that neuropsychiatric issues are under-recognized in this metabolic disease. Oculomotor disturbances, likely involving cerebellar motor and dorsal attention domains, have also been recently detailed by Stephen et al. [12]. In summary, LOGG is an ultra-rare degenerative neurological disease not only associated with motor impairments, but psychiatric and cognitive impairments as well [10].

Despite these complex characteristics, comprehensive population-level neuroimaging studies in LOGG have been scant. Fukumizu et al. [21] found low cerebral white matter density in 4 LOGG patients using computed tomography, while Jamrozik et al. [22] reported glucose hypometabolism in bilateral temporal and occipital lobes using positron emission tomography. One study using magnetic resonance spectroscopy reported lower N-acetylaspartate (NAA) in LOGG patients in the thalamus and occipital white matter [23]. Our recent study on the same cohort found lower NAA and higher myo-inositol in the cerebellum [24]. Grosso et al.’s MRI study [25] involving early, juvenile and late-onset forms of Tay-Sachs disease revealed basal ganglia and thalamic atrophy in early Tay-Sachs, cortical atrophy in juvenile Tay-Sachs, and cerebellar atrophy in LOTS, indicating that the neurobiology of LOTS differs from other forms of Tay-Sachs disease. Cerebellar atrophy has been the most consistent finding of the few LOGG imaging studies conducted thus far [26] [27] [22] [28] [29] [30] (including in our data [24]), but atrophy was not found to be associated with clinical variables (symptom severity, disease duration, etc.), hinting that deeper mechanistic insights beyond cerebellar atrophy are needed to better understand this disease. Other infrequent imaging findings in LOGG have included cerebral atrophy [31], thalamic hypodensities [31], mild midbrain and brainstem atrophy [32], and white matter lesions [33].

While the above findings represent some advancements in imaging the structural and metabolic impairments of LOGG, to the best of our knowledge, there have been no functional neuroimaging studies to date (either fMRI or other modalities) that have investigated LOGG or other forms of GM2 gangliosidosis. Thus, we targeted this gap herein using whole-brain fMRI data obtained from LOGG patients and matched healthy controls. Given the lack of LOGG fMRI studies and the challenge of developing robust hypotheses from other modalities owing to insufficient and inconclusive information, we did not develop specific regional hypotheses but undertook exploratory analyses with various fMRI analysis techniques (low-frequency fMRI signal strength, neurovascular coupling, static and time-varying functional connectivity, and network topology). As the first fMRI study in this disease cluster, this multi-pronged approached helped us obtain the landscape of functional impairments in LOGG, which will aid in developing hypothesis-driven studies in the future. We also probed the association of significant imaging measures with clinical variables and gene expression to understand the factors influencing functional impairments in LOGG.

## 2. Methods

### 2.1. Participants

Informed consent was obtained from all participants and the study was approved by the Mass General Brigham (formerly Partners HealthCare) Institutional Review Board (IRB). Seven adults with a genetically confirmed LOGG diagnosis were recruited either through the Leukodystrophy Clinic at the Massachusetts General Hospital or the National Tay-Sachs and Allied Diseases Association (NTSAD). The diagnosis was defined through near-absence (or absence) of HEX enzymatic activity (in serum or white blood cells), or mutation analysis of *HEXA* and *HEXB* genes. Across patients, five were diagnosed with the LOTS subtype of LOGG and two with the LOSD subtype. These subtypes have established differences [24], but we could not probe them within our cohort because of the sample sizes. Since LOGG is an ultra-rare disease, our sample size was understandably small but still respectable considering its prevalence of 1 in ∼300,000 in the general population. All LOGG patients were examined by a board-certified neurologist (C.D.S., ataxia specialist). Seven age- and sex-matched healthy controls were carefully screened and recruited. Inclusion criteria were age ≥18 years, able and willing to undergo MRI, and able and willing to provide informed consent.

The following clinical/behavioral measures were assessed [12]: Clinical ataxia rating scales: Brief Ataxia Rating Scale (BARS) [34], Scale for the Assessment and Rating of Ataxia (SARA) [35], Friedreich Ataxia Rating Scale (FARS) [36], and LOTS severity scale [37]; Cerebellar Neuropsychiatric Rating Scale (CNRS), a care-giver reported measure of psychiatric symptoms [38]; Cerebellar Cognitive Affective Syndrome Scale (CCAS), a specially designed cognitive rating scale to assess cognitive dysfunction seen in cerebellar disease [39]; sleep scales: Pittsburgh Sleep Quality Index (PSQI) [40] and Epworth sleepiness scale [41]; and a scale measuring dysphagia, the Eating Assessment Tool (EAT10) [42]. As previously described, SARA, FARS and BARS scores were adjusted for clinical weakness to allow better assessment of ataxia severity [12].

### 2.2. MRI data acquisition

BOLD fMRI data were acquired in a 3T Siemens Trio MRI scanner (Siemens Healthcare, Erlangen, Germany). The scan parameters were as follows: 2D echo planar imaging (EPI) sequence, TR = 2000 ms, TE = 30 ms, flip angle = 90°, voxel size = 2.5×2.5×4 mm^3^, field of view = 200×200 mm, scan duration = 10 min, 36 slices, with whole-brain coverage. High-resolution T1-weighted anatomical images were acquired with the following parameters: 3D magnetization-prepared rapid acquisition of gradient echo (MPRAGE) sequence: TR = 2530 ms, TE = 1.64 ms, flip angle = 7°, TI = 1200 ms, voxel size = 1×1×1 mm^3^, field of view = 256×256 mm.

### 2.3. fMRI data pre-processing

All analyses were performed in MATLAB R2019b, with pre-processing done using CONN v18b [43], based on SPM12 [44]. The following steps were performed: realignment, co-registration to MNI space, outlier detection using ARtifact detection Tools (ART) [45] (including identifying high-motion volumes with motion over half the voxel size), and denoising. Denoising was performed by regressing out the following nuisance covariates: top 10 principal components of white matter and CSF signals, 12 head motion parameters (3 translation, 3 rotation and their derivatives), and outlier volumes identified using ART. Thereafter, linear detrending and high-pass filtering (0.01Hz) were performed. The largest permitted head motion was half the voxel size (1.25mm); no participants were excluded. Aggregate head motion (mean framewise displacement [46]) was not statistically different between the groups (*p*=0.29). Spatial smoothing was not performed because we did not conduct any voxel-level analyses, and fMRI time series were averaged anyway within regions of interest (ROIs) (explained later). Bandpass filtering was performed at a later stage in the analysis pipeline (see section 2.7) because some of the analyses required unfiltered data. Visual inspection was performed at every stage of processing to ensure that accurate and reliable outputs were generated. Of particular importance, co-registrations and segmentations of the cerebellum in LOGG were visually inspected to ensure correctness.

The fMRI signal is a convolution of the latent neural activity and the hemodynamic response function (HRF), where the HRF represents mechanisms occurring between neuronal firing and the corresponding BOLD response. The HRF is found to vary across the brain and across individuals [47]. This HRF variability confounds fMRI connectivity estimates [48] [49] [50]. Additionally, group differences in connectivity between healthy and disease groups are confounded by HRF variability [8] [51]. Hence, deconvolution was carried out on fMRI data [52] to estimate the HRF and obtain deconvolved (HRF variability minimized) fMRI data, like in several recent studies [53] [54] [55] [56] [57]. Deconvolved fMRI data was used in all further analyses.

Given the high dimensionality of whole-brain 3D+time fMRI data, a functional brain parcellation was used for dimensionality reduction with 272 homogeneous ROIs, comprising of 242 cortical ROIs defined by the Power atlas [5], 16 subcortical ROIs defined by the Harvard-Oxford atlas [58], and 14 cerebellar ROIs defined by the Buckner atlas [59]. Mean time series was computed for each ROI by averaging data from all gray matter voxels within the ROI (50% gray matter mask), which were then used in further processing.

### 2.4. Analysis framework

The fMRI literature describes various techniques to infer clinically relevant information from fMRI data; we employed some of the most commonly used ones herein. Contrary to many studies that focus on just one technique, we used multiple techniques as this is the first fMRI study in this disease cluster. We first gathered observations using each technique and later interpreted the findings as a convergence of multiple observations, to aid hypothesis-driven studies in the future. We briefly introduce each of these approaches as well as their rationale.

#### (i) Low-frequency BOLD fMRI signal strength

BOLD activation has been commonplace with task fMRI, and its analogue with resting-state fMRI is called the fractional amplitude of low frequency fluctuations (fALFF) [7]. FALFF quantifies the magnitude of low frequency BOLD fluctuations (0.01–0.10 Hz) relative to the entire frequency range, and is found to be impaired in neurological disorders such as Parkinson’s disease [60] and multiple sclerosis [61]. This represents average neuronal activity and is a reasonably popular approach (PubMed: 1000+ papers until 2021, 82% of them since 2015). We assessed fALFF impairments in LOGG, consistent with studies in other neurological disorders [60] [61].

#### (ii) Neurovascular coupling

While fALFF measures BOLD signal strength, it does not untangle neuronal signal and neurovascular coupling (HRF) that form the BOLD signal. Typically, HRF is considered a confound in fMRI analysis since neural activity, not neurovascular coupling, is of interest in most studies. As described earlier, HRF variability confounds connectivity. However, the study of neurovascular coupling is of interest to the community. For instance, elevated strength and reduced latency of HRF was observed in posttraumatic stress disorder [8], while reduced strength and latency were observed in autism spectrum disorder [51] and obsessive-compulsive disorder [62]. There have been no HRF studies on neurological disorders, although underlying neurochemical and extracellular mechanisms that modulate HRF are impacted by the degenerative nature of such diseases. Here, we assessed HRF impairments in LOGG, with the purpose of deriving unique and complementary information that augments observations from other analyses performed in this study. Interestingly, genes have been found to influence the HRF by 24–51% [63], further suggesting its relevance for study in LOGG (a genetic disease).

#### (iii) Static and dynamic functional connectivity

While fALFF and HRF quantify regional properties, they do not measure how the different brain regions communicate with each other. Functional connectivity (FC) quantifies co-activation between pairs of brain regions, and is sensitive to psychiatric [64] [56] and neurological [1] [2] [3] [60] disease pathology. Static FC (SFC) measures the strength of connectivity between regions and has been extensively utilized (PubMed: 11,000+ papers until 2021, 72% of them since 2015). We studied whole-brain SFC impairments in LOGG.

SFC provides one value per connection for the entire scan duration spanning several minutes, which does not capture temporal variations in connectivity within time scales of seconds to minutes. For this reason, although SFC remains widely used, the study of dynamic FC (DFC) [65] has gained considerable traction, especially in the past 5 years (PubMed: 1500+ papers until 2021, 78% since 2015). DFC provides fundamentally different information from SFC [66] [65]. DFC is sensitive to pathology [67] [68], is related to real world cognitive behaviors [69], and may be more useful as a biomarker than SFC [70] [71]. Reduced temporal variance of DFC (vDFC) is associated with psychiatric illnesses as well as lower behavioral performance in healthy people [56] [70] [72], that is, inability to dynamically adjust (thoughts and behaviors) to changing body and environmental conditions. We sought to identify whole-brain vDFC reduction in LOGG.

#### (iv) Network topology of the SFC network

fALFF and HRF characterize regional properties, and at the next level of complexity, FC characterizes communication between pairs of regions in isolation. However, they do not describe the nature of the ensemble of thousands of connections. Complex network modeling using graph analysis methods can quantify network topology [6]. This approach uses the pattern in which connections coexist to make inferences about network structure. Graph measures are sensitive to psychiatric and neurological disease pathology [1] [73] [74]. Much like HRF and DFC, this is an emerging technique (PubMed: 1800+ papers until 2021, 74% since 2015). Along with fALFF, HRF, SFC and DFC, we sought to identify impairments in graph measures in LOGG. Given this description of our analysis framework, we will next describe the implementation of each technique.

### 2.5. Measuring impairments in low frequency BOLD signal strength using fALFF

FALFF was computed by applying Fourier transform to the ROI time series and calculating a ratio in the frequency domain. Specifically, fALFF is the ratio of low-frequency fMRI signal power (0.01–0.1 Hz) to the total power in entire frequency range (0–0.25 Hz for our data with TR=2s) [75]. Since neuronal signal correlates of fMRI reside mostly in this low frequency band [5], and signal outside this band (<0.01Hz and >0.1Hz) is generally considered noise, this ratio measures BOLD ‘activation’ at rest. FALFF was computed for each of the 272 ROIs, and group comparisons were performed (*p*<0.05, FDR corrected). All statistical tests in the entire study were controlled for head motion (mean framewise displacement [46]); this will not be repeatedly mentioned elsewhere.

### 2.6. Measuring neurovascular coupling impairments using the HRF

HRF shape can be characterized by three parameters: response height (RH), time-to-peak (TTP), and full-width-at-half-maximum (FWHM) [47] (refer to Fig.1 in [48]). RH is the HRF amplitude, TTP represents the latency of BOLD response, and FWHM is related to BOLD response duration. Given that TTP has a typical range of 2.5–6.5s and FWHM has a typical range of 1–2s [47], our data with TR=2s does not have satisfactory temporal resolution. Thus, only RH was examined here. The deconvolution step (described before) provided us the HRF parameters in the 3D MNI space. Mean RH values were extracted from the 272-ROI atlas mentioned earlier, providing us 272 RH values per subject. A group comparison was then performed (*p*<0.05, FDR corrected) to measure RH impairments in LOGG.

**Figure 1.**
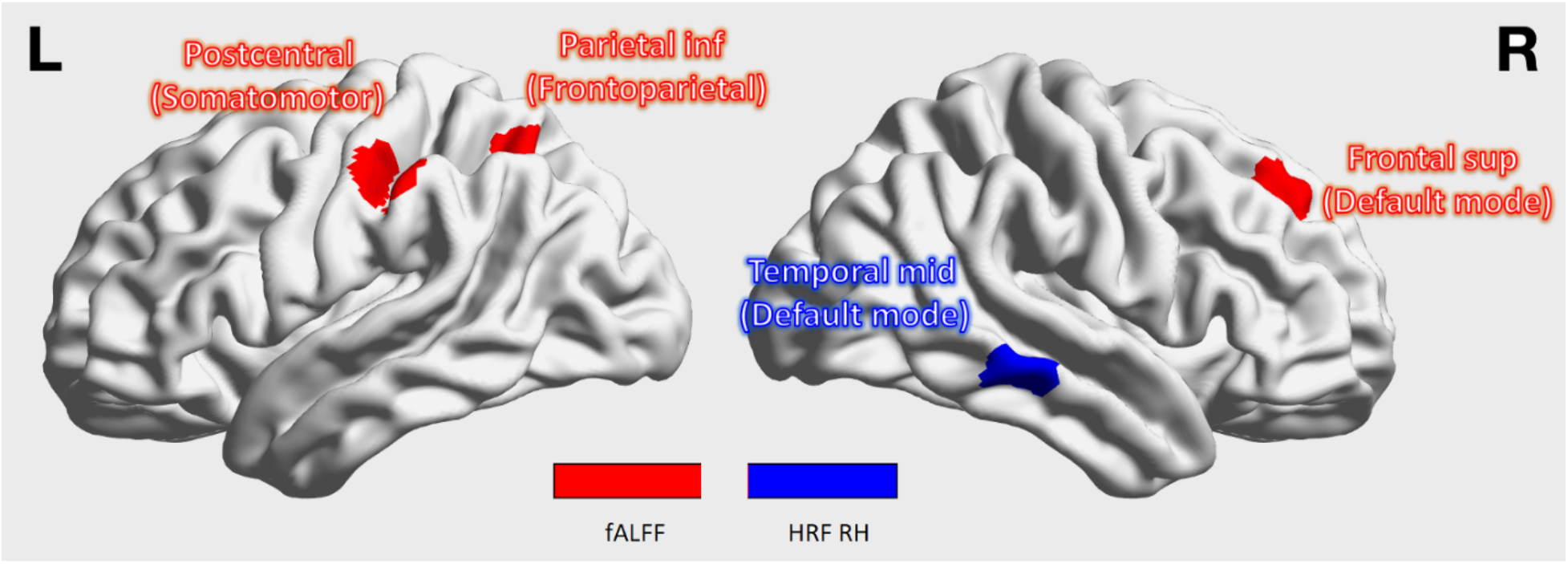
fALFF (red) and HRF response height (RH) (blue) results. fALFF was reduced and RH was elevated in LOGG, implying reduced BOLD signal strength and elevated neurovascular coupling strength in the respective regions. In the figure, both region names and corresponding network assignments are shown, with the network mentioned within parentheses. This convention also applies to Figures 2 and 3. The visualizations in Figures 1, 2 and 3 were generated using BrainNet Viewer [85]. Inf = inferior; sup = superior.

### 2.7. Static and dynamic functional connectivity modeling

Bandpass filtering (0.01–0.1Hz) was performed on ROI time series data prior to connectivity modeling, using a 15^th^ order finite impulse response filter. SFC was computed using Pearson’s product-moment correlation coefficient, giving us a 272×272 connectivity matrix per subject. Group differences in SFC are reported (*p*<0.05, FDR corrected).

To compute DFC, a sliding window Pearson’s correlation approach was employed (see [76] for a thorough review of DFC methods). To avoid arbitrarily choosing the window length, a variable window length approach was used [70] to determine the window length adaptively based on the augmented Dickey-Fuller (ADF) test (quantifies time series stationarity). This technique searched for an optimal minimum window length (between 40s and 60s [76]) that preserves stationarity. Although Hutchison et al. [76] recommend choosing the window length within the range of 30−60s, 40s was chosen to ensure at least 20 data points (because TR=2s) to be able to reliably estimate a correlation [77]. Then, to quantify variability of connectivity over time, temporal variance of the DFC time series (vDFC) was computed to obtain a 272×272 vDFC matrix per subject. This approach has been popularized by several recent studies [56] [78] [70] [79]. Whole-brain group differences in vDFC were assessed (*p*<0.05, FDR corrected).

### 2.8. Graph analysis using static functional connectivity networks

To measure aberrations in brain network topology that cannot be ascertained using bivariate FC alone, complex network modeling was performed using graph measures. Specifically, we studied the pathological shift in balance between functional segregation and integration in LOGG. Graph measures were computed using the 272×272 SFC network in each subject. Each SFC value (across subjects) was statistically compared against null (FC=0), and all non-significant connections (*p*>0.05) were forced to 0. Although we refer the readers to Rubinov and Sporns [6] for a detailed theoretical and mathematical rendering of these concepts, we briefly explain the measures here for the benefit of the readers. Segregation (one value per ROI) was measured independently using the following three graph measures.

i. Clustering coefficient – fraction of each ROI’s neighboring connections that are neighbors of each other (measures dense-connectedness; higher value implies higher functional segregation or elevated local processing of functionally specialized subnetworks).
ii. Local efficiency – average of inverse shortest path lengths (i.e., reachability) between the given ROI and all other regions (higher value implies that other brain regions find it easier to communicate with the given ROI, i.e., higher segregation).
iii. Node strength – average connectivity value of all the connections associated with the given ROI (higher value implies overall higher connectivity between the ROI and rest of the brain). Functional integration (one value per connection) was measured independently using the following two measures.
iv. Shortest path length – length of the functional communication channel between pairs of ROIs (higher length implies that it is more difficult for the two regions to communicate with each other, i.e., poorer functional integration).
v. Edge betweenness – number of all shortest paths in the whole network that contain the given connection (higher value implies that the given connection is important for the rest of the brain connectome, i.e., higher integration). It is restricted to integer values.

Network topological properties quantified by these measures are fundamentally different from bivariate SFC, and this study aimed to employ these complementary approaches to develop a multi-dimensional understanding of LOGG pathology. Group differences in each of these graph measures are reported (*p*<0.05, FDR corrected).

### 2.9. Associations between fMRI measures and non-imaging data (gene expression, clinical variables)

The Allen human brain atlas of gene expression [80] has been mapped to [81] the Desikan-Killiany (DK) whole-brain parcellation (FreeSurfer) with 62 regions [82], enabling the exploration of associations between imaging measures and gene expression. Traditionally, gene expression is derived from blood samples in the same individual and correlated with imaging values; however, in our analysis the Allen brain gene expression atlas provided us the spatial distribution of gene expressions across the brain in a normative healthy sample (taking years to develop [80]), which was correlated with the spatial distribution of imaging measures. Given that LOGG is a genetic disorder [12], we probed the associations between the DK parcellation gene expression map and the maps of regional fMRI measures in the DK parcellation space (fALFF, HRF RH, and segregation measures). As stated before, LOGG is known to be caused by single gene mutations [13], with LOTS due to mutations in *HEXA* and LOSD due to mutations in *HEXB*.

We first computed mean imaging values among all voxels within each DK parcellation ROI, giving us one value for each DK-ROI in each subject. We then computed fALFF, HRF RH, and segregation graph measures for each ROI, giving us a spatial distribution of these imaging measures across the brain. These maps were then averaged across subjects to obtain one spatial map for LOTS and another map for LOSD (separately for each measure). Finally, by vectorizing the 3D maps into 1D vectors and correlating them, we examined the association (*p*<0.05, FDR corrected) between imaging maps and gene expression maps (*HEXA* in LOTS and *HEXB* in LOSD), revealing the variance of HEX genetic information in our imaging findings. Such an approach has been adopted by imaging studies recently [83] [84]. We also assessed the associations between sixteen clinical/behavioral measures (mentioned earlier) and significant fMRI observations (significant fALFF, HRF, connectivity, and graph measures) (*p*<0.05, FDR corrected).

## 3. Results

**Table 1** provides participant demographics and clinical scores (thoroughly presented in an earlier study on the same cohort [12]). Age was not significantly different between groups (*p* = 0.95, *Z* = −0.06, Wilcoxon rank-sum test).

**Table 1.** Demographics and clinical variables. Other than age and sex, all remaining measures pertain to the LOGG group alone.

| Variable | mean ± SD | Range |
| --- | --- | --- |
| <u>Disease demographics</u> |  |  |
| Age (LOGG) (years) | 42.9 ± 7.4 | 22 – 62 |
| Sex (LOGG) | 4 M, 3 F | - |
| Age (control) (years) | 42.7 ± 7.2 | 23 – 62 |
| Sex (control) | 4 M, 3 F | - |
| Age at disease onset | 15 ± 3.6 | 8 – 26 |
| Disease duration (y) | 27.9 ± 4.5 | 12 – 39 |
| <u>Clinical rating scales</u> |  |  |
| BARS | 5.4 ± 2.3 | 1.5 – 15 |
| SARA | 10.4 ± 1.9 | 4.5 – 16.5 |
| FARS | 33.1 ± 6.5 | 18 – 55.5 |
| LOGG severity score | 7.1 ± 1.5 | 4 – 13 |
| <u>Cognitive/psychiatric scales</u> |  |  |
| CCAS | 98.9 ± 5.9 | 76 – 114 |
| CNRS (total) | 24.9 ± 6.1 | 6 – 43 |
| Attentional control <sup>a</sup> | 7.7 ± 1.5 | 2 – 11 |
| Emotional control <sup>a</sup> | 5.4 ± 1.2 | 2 – 9 |
| Social skillset <sup>a</sup> | 6 ± 2.6 | 2 – 16 |
| Autism spectrum <sup>a</sup> | 2.1 ± 1.4 | 0 – 8 |
| Psychosis spectrum <sup>a</sup> | 3.7 ± 1.6 | 0 – 10 |
| <u>Sleep scales</u> |  |  |
| PSQI | 7.9 ± 3.2 | 2 – 20 |
| ESS | 9.7 ± 3.1 | 2 – 21 |
| <u>Other scales</u> |  |  |
| EAT10 | 9 ± 4.3 | 0 – 22 |

<sup>a</sup> These are subscales of CNRS.
LOGG = late-onset GM2 gangliosidosis; CNRS = cerebellar neuropsychiatric rating scale; CCAS = cerebellar cognitive affective syndrome scale; FARS = Friedreich ataxia rating scale; BARS = brief ataxia rating scale; SARA = Scale for the Assessment and Rating of Ataxia; EAT10 = eating assessment tool; PSQI = Pittsburgh sleep quality index; ESS = Epworth Sleepiness Scale.

### 3.1. BOLD signal strength (fALFF) and neurovascular coupling (HRF) results

Overall hypo-activity was observed in LOGG as fALFF was lower in LOGG vs. controls in somatosensory and frontoparietal regions (somatomotor, frontoparietal and default mode networks [DMN]). On the other hand, HRF RH was higher in LOGG in the middle temporal (DMN) region (**Figure 1, Figure 4, Table 2**).

**Table 2.** fALFF and HRF results (whole-brain LOGG vs. control). FALFF was reduced in LOGG in all three regions. HRF response height was elevated in LOGG in the one region. Inf = inferior; sup = superior; mid = middle.

|  | Network | Region / node | MNI centroid | p-value | T statistic <sup>a</sup> | Effect size |
| --- | --- | --- | --- | --- | --- | --- |
| **fALFF: LOGG vs. control** |  |  |  |  |  |  |
| **1** | Somatomotor | Postcentral L | (-54, -23, 43) | 0.0007 | -4.41 | 2.34 |
| **2** | Default mode | Frontal Sup R | (22, 39, 39) | 0.0003 | -4.88 | 2.26 |
| **3** | Frontoparietal task control | Parietal Inf L | (-28, -58, 48) | 0.0049 | -3.38 | 1.99 |
| **HRF response height: LOGG vs. control** |  |  |  |  |  |  |
| **1** | Default mode | Temporal Mid R | (65, -31, -9) | 0.0096 | 3.03 | 1.59 |

<sup>a</sup> Positive *T* statistic: Higher in LOGG compared to controls; negative *T* statistic: lower in LOGG.

### 3.2. Static functional connectivity (SFC) results

Overall hyper-connectivity was observed in LOGG with all four identified connections (**Figure 2, Figure 4, Table 3**). With an uncorrected threshold of *p*<0.05, 62.9% of all SFC values were higher in LOGG; with *p*<0.001 uncorrected, 73% were higher in LOGG, corroborating the pattern observed with corrected statistics. The four connections were mainly associated with cognitive control networks (attention, task control, salience), primarily in prefrontal and lateral parietal regions. Half the connections were within the attention networks.

**Figure 2.**
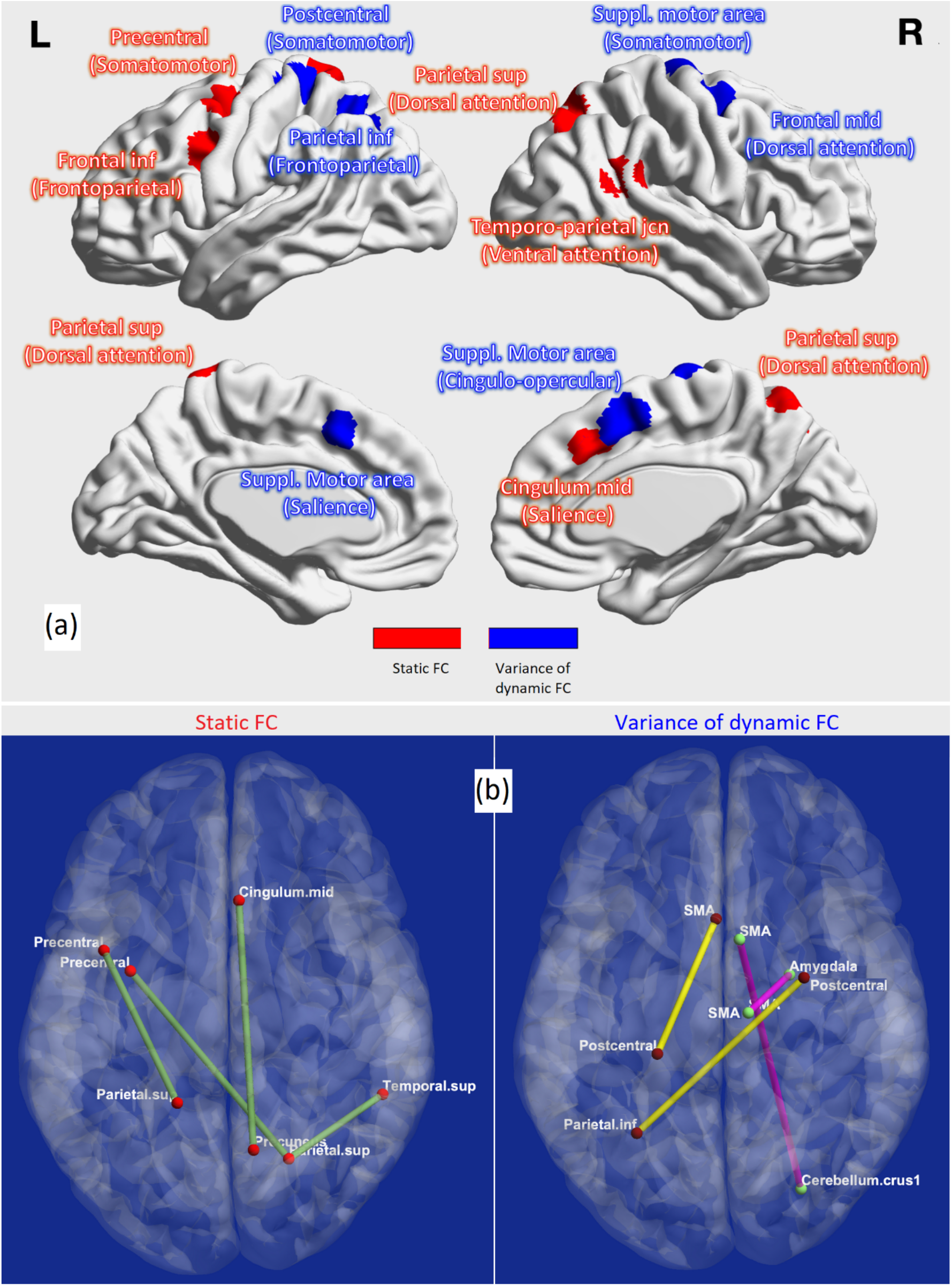
Functional connectivity results. (a) Regions associated with static FC (red) and variance of dynamic FC (blue) impairments in LOGG. (b) Impaired connections in LOGG. Bottom-left: elevated static FC impairments are shown in green; bottom-right: reduced dynamic FC impairments are shown in yellow and elevated dynamic FC impairments are shown in pink. Sup = superior; inf = inferior; mid = middle; SMA = supplementary motor area; jcn = junction.

**Figure 3.**
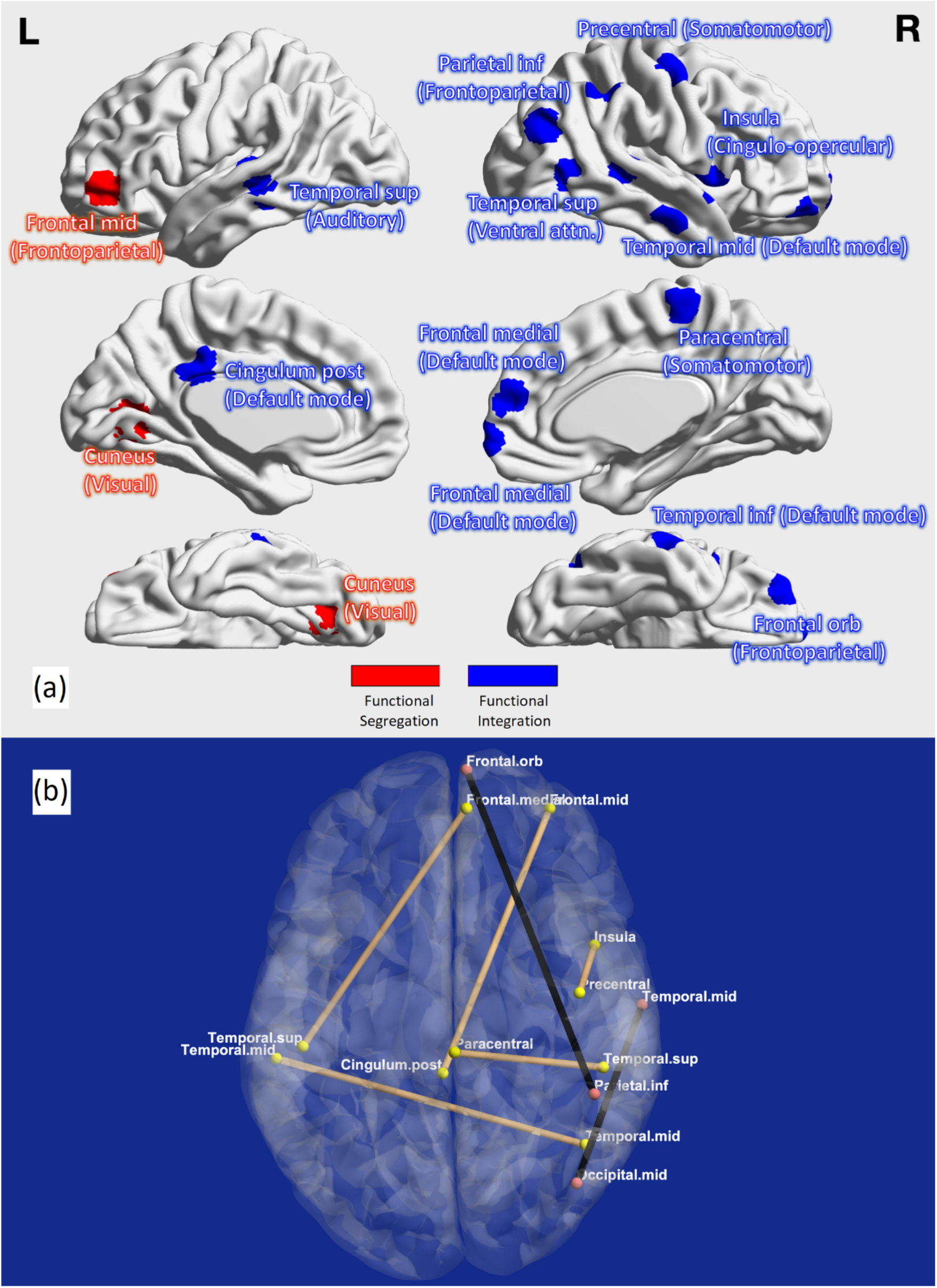
Graph analysis results. (a) Regions associated with functional segregation (red) and functional integration (blue) impairments in LOGG. (b) Connections with impaired integration in LOGG: connections with reduced edge betweenness are shown in brown, connections with elevated shortest path length are shown in black. Prominent right lateralization was observed. Orb = orbital; med = medial; mid = middle; sup = superior; inf = inferior; post = posterior.

**Figure 4.**
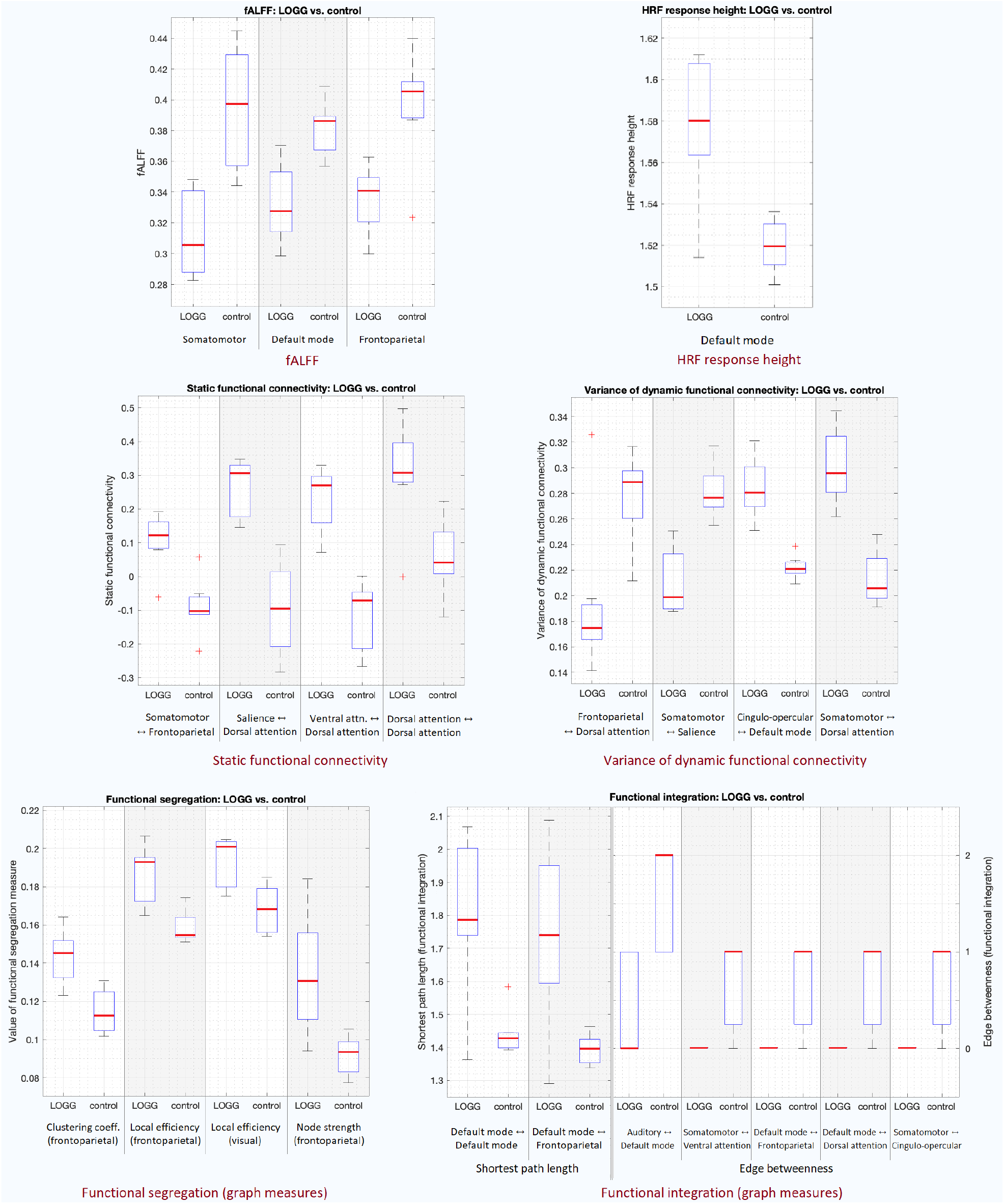
Boxplots of all presented results. Top row: fALFF (left) and HRF RH (right). Middle row: SFC (left) and vDFC (right). Bottom row: segregation (left) and integration (right). The red line represents the median, the box extends from 25^th^ to 75^th^ quartile, the dotted lines extend from 1^st^ to 99^th^ quartile, and the rare ‘+’ markings represent the outliers.

**Table 3.** Static functional connectivity results (whole-brain LOGG vs. control). LOGG group exhibited hyperconnectivity; all four connections, mainly associated with attention and task control networks, were elevated in the disease. Inf = inferior; sup = superior; junc = junction.

|  | Network connectivity | Connection | MNI centroids | p-value | T stat <sup>a</sup> | Effect size |
| --- | --- | --- | --- | --- | --- | --- |
| 1 | Somatomotor ↔<br>Frontoparietal task control | Precentral L ↔<br>Frontal Inf L | (-16, -46, 73) ↔<br>(-41, 6, 33) | $9.5 \times 10^{-5}$ | 5.54 | 3.06 |
| 2 | Salience ↔<br>Dorsal attention | Cingulum Mid R<br>↔ Parietal Sup R | (5, 23, 37) ↔<br>(10, -62, 61) | $6.9 \times 10^{-5}$ | 5.73 | 3.17 |
| 3 | Ventral attention ↔<br>Dorsal attention | Temporo-parietal<br>Junc. R ↔<br>Parietal Sup R | (54, -43, 22) ↔<br>(22, -65, 48) | $9.7 \times 10^{-5}$ | 5.53 | 3.05 |
| 4 | Dorsal attention ↔<br>Dorsal attention | Frontal Mid R ↔<br>Parietal Sup L | (22, -65, 48) ↔<br>(-32, -1, 54) | $1.1 \times 10^{-4}$ | 5.45 | 3.02 |

<sup>a</sup> Positive T statistic: Higher connectivity in LOGG compared to controls; negative T statistic: lower in LOGG.

### 3.3. Dynamic functional connectivity (vDFC) results

Four connections were identified with impaired vDFC in LOGG (**Figure 2, Figure 4, Table 4**) associated with cognitive control (attention, task control, salience), DMN and somatomotor networks, primarily in supplementary motor and lateral prefrontal regions. Contrary to our expectation of finding lower variability of connectivity, two connections exhibited lower vDFC while two others showed higher vDFC in LOGG. However, with an uncorrected threshold of *p*<0.05, 61% of all vDFC values were lower in LOGG; with *p*<0.001 uncorrected, 67.7% were lower in LOGG. This implied that although only half of the identified significant connections had lower vDFC in LOGG, a general pattern of lower vDFC was noticeable by examining the numbers with uncorrected stats, corroborating previous vDFC studies [56]. Because of the specific involvement of the cerebellum in LOGG pathology [24], SFC and vDFC impairments only within the cerebellum were additionally assessed as a tertiary analysis (see Supplemental Information section S1).

**Table 4.** Dynamic functional connectivity results (whole-brain LOGG vs. control). LOGG group exhibited both reduced and elevated variability of connectivity. Three of four connections involved the supplementary motor area, and two connections involved the middle frontal gyrus. Inf = inferior; mid = middle; suppl = supplementary.

|  | Network connectivity | Connection | MNI centroids | p-value | T stat <sup>a</sup> | Effect size |
| --- | --- | --- | --- | --- | --- | --- |
| **1** | Frontoparietal task control ↔ Dorsal attention | Parietal Inf L ↔ Frontal Mid R | (-28, -58, 48) ↔ (29, -5, 54) | $1.3 \times 10^{-5}$ | -6.78 | 3.64 |
| **2** | Somatomotor ↔ Salience | Postcentral L ↔ Suppl. Motor Area L | (-21, -31, 61) ↔ (-1, 15, 44) | $9.9 \times 10^{-5}$ | -5.52 | 3.01 |
| **3** | Cingulo-opercular task control ↔ Default mode | Suppl. Motor Area R ↔ Cerebellum Crus1 R | (7, 8, 51) ↔ (28, -77, -32) | $6.6 \times 10^{-5}$ | 5.76 | 3.12 |
| **4** | Somatomotor ↔ Dorsal attention | Suppl. Motor Area R ↔ Frontal Mid R | (10, -17, 74) ↔ (24, -4, -18) | $9.8 \times 10^{-5}$ | 5.52 | 3.01 |

<sup>a</sup> Positive *T* statistic: Higher variability of connectivity in LOGG vs. controls; negative *T* statistic: lower in LOGG.

### 3.4. Graph analysis results

Functional segregation was significantly higher in LOGG across the board (**Figure 3, Figure 4, Table 5**) in lateral prefrontal and visual regions. Specialized densely connected local processing is especially predominant in prefrontal and visual regions of the healthy brain [64], the former for cognitive control and the latter for visual perception. With an uncorrected threshold of *p*<0.05, 100% of all segregation measures were higher in LOGG.

**Table 5.**
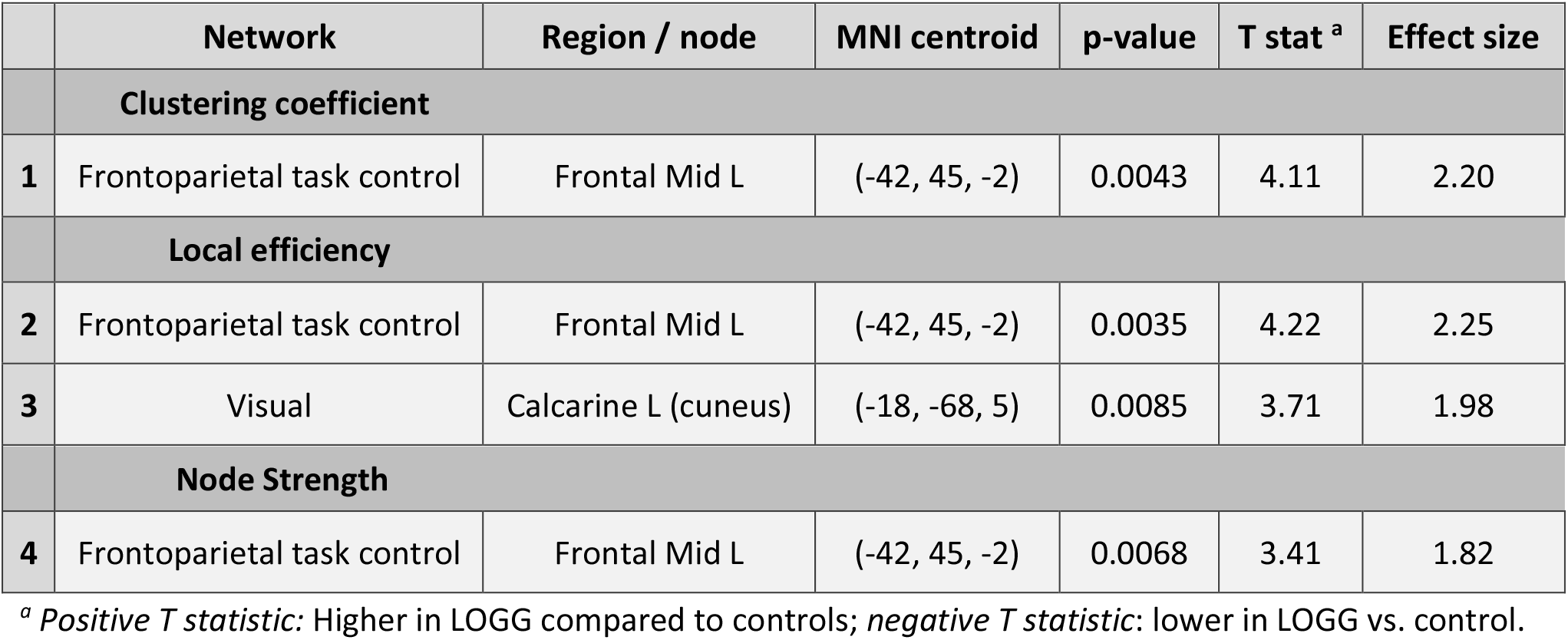
Functional segregation (graph analysis) results (whole-brain LOGG vs. control). LOGG group exhibited elevated segregation in task control and visual regions, implying abnormally heightened processing within specialized prefrontal and visual subnetworks. Mid = middle

|  | Network | Region / node | MNI centroid | p-value | T stat <sup>a</sup> | Effect size |
| --- | --- | --- | --- | --- | --- | --- |
|  | **Clustering coefficient** |  |  |  |  |  |
| **1** | Frontoparietal task control | Frontal Mid L | (-42, 45, -2) | 0.0043 | 4.11 | 2.20 |
|  | **Local efficiency** |  |  |  |  |  |
| **2** | Frontoparietal task control | Frontal Mid L | (-42, 45, -2) | 0.0035 | 4.22 | 2.25 |
| **3** | Visual | Calcarine L (cuneus) | (-18, -68, 5) | 0.0085 | 3.71 | 1.98 |
|  | **Node Strength** |  |  |  |  |  |
| **4** | Frontoparietal task control | Frontal Mid L | (-42, 45, -2) | 0.0068 | 3.41 | 1.82 |

<sup>a</sup> Positive *T* statistic: Higher in LOGG compared to controls; negative *T* statistic: lower in LOGG vs. control.

Functional integration was significantly lower in LOGG (**Figure 3, Figure 4, Table 6**) in cognitive control (attention, task control), DMN, and somatomotor networks. Five of these seven connections involved the DMN (mainly connected to control/sensory networks). With an uncorrected threshold of *p*<0.05, 55% of all values indicated lower integration in LOGG. Lastly, upon correlating all significant imaging findings across the board with age, we found that none of the measures were influenced by age (*p*>0.05).

**Table 6.** Functional integration (graph analysis) results (whole-brain LOGG vs. control). LOGG group exhibited reduced integration. All connections, mainly associated with default mode, attention, and task control networks, were associated with poorer integration in LOGG. Higher shortest path length implies poorer integration, so does lower edge betweenness. Inf = inferior; sup = superior; mid = middle; post = posterior; ctrl = control.

|  | Network connectivity | Connection | MNI centroids | p-value | Statistic <sup>a</sup> | Effect size |
| --- | --- | --- | --- | --- | --- | --- |
| **Shortest path length** (T statistic reported from Student's T test) |  |  |  |  |  |  |
| **1** | Default mode ↔<br>Default mode | Temporal Mid R ↔<br>Temporal Inf R | (43, -72, 28) ↔<br>(65, -12, -19) | $2.2 \times 10^{-6}$ | 8.01 | 4.27 |
| **2** | Default mode ↔<br>Frontoparietal task control | Frontal Medial R ↔<br>Parietal Inf R | (6, 67, -4) ↔<br>(49, -42, 45) | $3.8 \times 10^{-5}$ | 6.09 | 3.26 |
| **Edge betweenness centrality** (Z statistic reported from Wilcoxon rank-sum test) |  |  |  |  |  |  |
| **3** | Auditory ↔<br>Default mode | Temporal Sup L ↔<br>Frontal Medial R | (-49, -26, 5) ↔<br>(6, 54, 16) | $1.4 \times 10^{-5}$ | -6.73 | 3.61 |
| **4** | Somatomotor ↔<br>Ventral attention | Paracentral R ↔<br>Temporal Sup R | (2, -28, 60) ↔<br>(52, -33, 8) | $4.8 \times 10^{-4}$ | -4.62 | 2.47 |
| **5** | Default mode ↔<br>Frontoparietal task control | Cingulum Post L ↔<br>Frontal Orb R | (-2, -35, 31) ↔<br>(34, 54, -13) | $5.3 \times 10^{-4}$ | -4.57 | 2.48 |
| **6** | Default mode ↔<br>Dorsal attention | Temporal Mid L ↔<br>Temporal Mid R | (-58, -30, -4) ↔<br>(46, -59, 4) | $5.3 \times 10^{-4}$ | -4.57 | 2.45 |
| **7** | Somatomotor ↔<br>Cingulo-opercular task ctrl | Precentral R ↔<br>Insula R | (44, -8, 57) ↔<br>(49, 8, -1) | $6.4 \times 10^{-4}$ | -4.46 | 2.38 |

<sup>a</sup> *Positive statistic:* Higher value in LOGG compared to controls; *negative statistic:* lower in LOGG.

### 3.5. Associations between fMRI measures and clinical variables

We found significant associations between certain LOGG imaging measures and clinical variables (**Figure 5, Table 7**). The figure shows LOSD patients in a different color, from which it appears that these associations were not driven by differences between LOTS and LOSD; however, we could not demonstrate this statistically because there were only two LOSD patients. Cognitive control networks were featured in 4 of the 6 associations. Among the clinical variables were neurocognitive measures, ataxia scales, disease duration, and age at disease onset. All associations followed the expected pattern of clinically worse imaging values relating to higher severity in clinical measures. Supplemental Information section S2 shows associations with subscales of certain clinical metrics.

**Figure 5.**
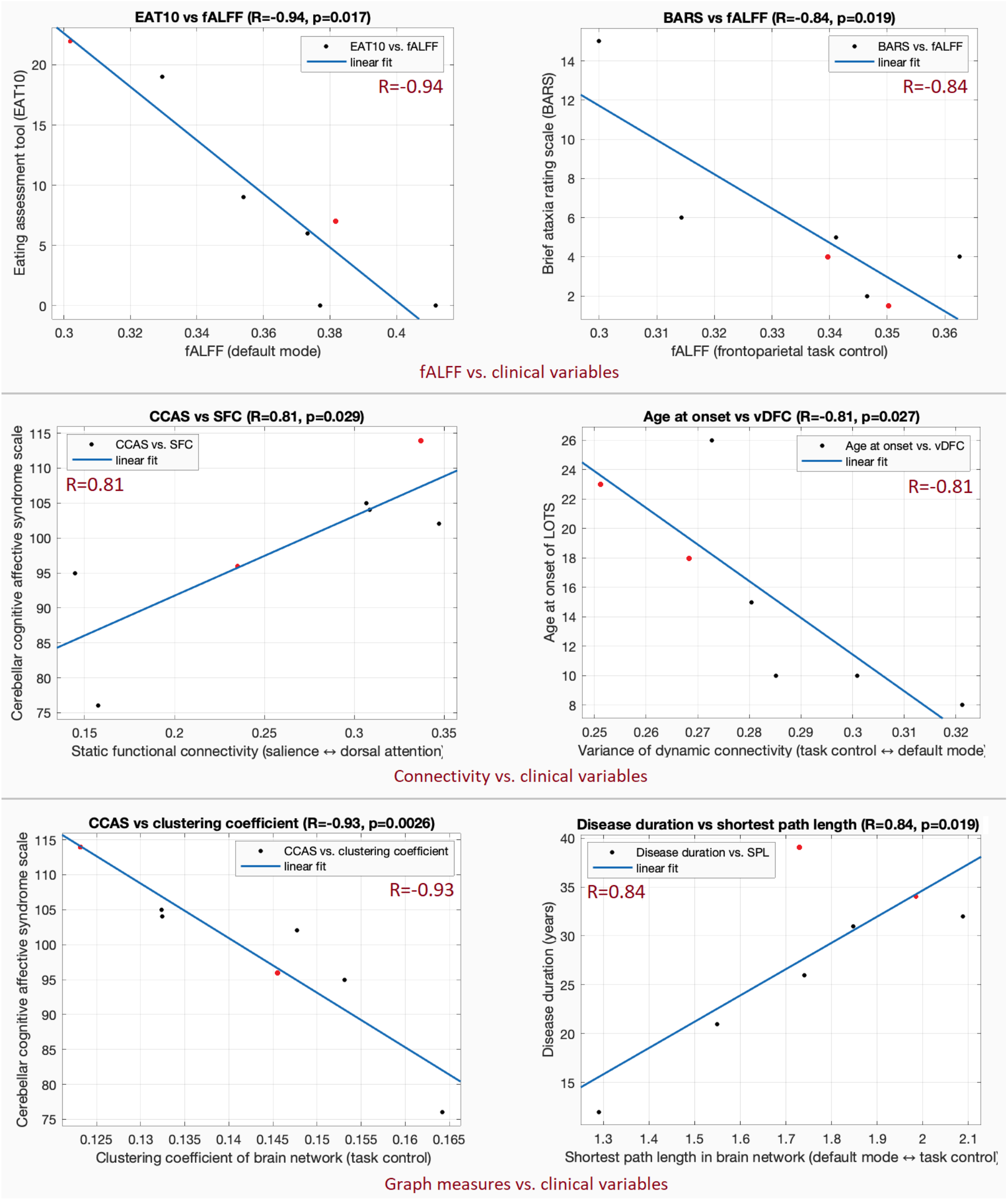
Associations between clinical variables and fMRI measures. Top row: fALFF vs. EAT10 (left, R=−0.94, p=0.002) and fALFF vs. BARS (right, R=−0.84, p= 0.019). Middle row: SFC vs. CCAS (left, R=0.81, p=0.029) and vDFC vs. age of disease onset (right, R=−0.81, p=0.027). Bottom row: segregation vs. CCAS (left, R=−0.93, p=0.003) and integration vs. disease duration (right, R=0.84, p=0.019). EAT10 = eating assessment tool, BARS = brief ataxia rating scale, CCAS = cerebellar cognitive affective syndrome scale. The two LOSD patients are shown as red points; it is apparent that the associations were not driven by differences in LOTS and LOSD.

**Table 7.** Significant associations between clinical variables and fMRI measures in LOGG.

|  | Clinical variable | Network | Connection / Node | R-value | R<sup>2</sup> value | p-value |
| --- | --- | --- | --- | --- | --- | --- |
| **fALFF** |  |  |  |  |  |  |
| 1 | EAT10 | Default mode | Frontal Medial R | −0.94 | 0.88 | 0.002 |
| 2 | BARS | Frontoparietal task control | Parietal Inf L | −0.84 | 0.7 | 0.019 |
| **Static functional connectivity** |  |  |  |  |  |  |
| 3 | CCAS | Salience ↔ Dorsal attention | Cingulum Mid R ↔<br>Parietal Sup R | 0.81 | 0.65 | 0.029 |
| **Variance of dynamic functional connectivity** |  |  |  |  |  |  |
| 4 | Age at disease onset | Cingulo-opercular task control<br>↔ Default mode | Suppl. Motor Area R<br>↔ Cerebellum Crus1 R | −0.81 | 0.66 | 0.027 |
| **Complex network measures** |  |  |  |  |  |  |
| 5 | CCAS | Clustering coefficient: fronto-<br>parietal task control | Frontal Mid L | −0.93 | 0.86 | 0.003 |
| 6 | Disease duration | SPL: Default mode ↔<br>Frontoparietal task control | Frontal Medial R ↔<br>Parietal Inf R | 0.84 | 0.7 | 0.019 |

EAT10 = eating assessment tool, BARS = brief ataxia rating scale, CCAS = cerebellar cognitive affective syndrome scale, SPL = shortest path length, sup = superior, inf = inferior, mid = middle, suppl = supplementary.

### 3.6. Associations between fMRI measures and gene expression

We found significant negative association between local efficiency and HEXB gene expression in LOSD patients (R=−0.55, p=4×10^−6^), but no significant association with *HEXA* in LOTS patients (R=−0.005, p=0.97) (**Figure 6**). HRF RH was significantly positively associated with both *HEXA* in LOTS (R=0.29, p=0.0225) and HEXB in LOSD (R=0.29, p=0.0229). These observations suggest a genetic basis for neurovascular coupling patterns in the brains of patients with LOTS and LOSD (about 8.5% variance in HRF RH explained by these genes), as well as a genetic basis for the ease of communication in the brains of those with LOSD (30.3% variance in local efficiency explained by HEXB in LOSD). **Figure 7** illustrates a flowchart of our entire analysis strategy and corresponding results.

**Figure 6.**
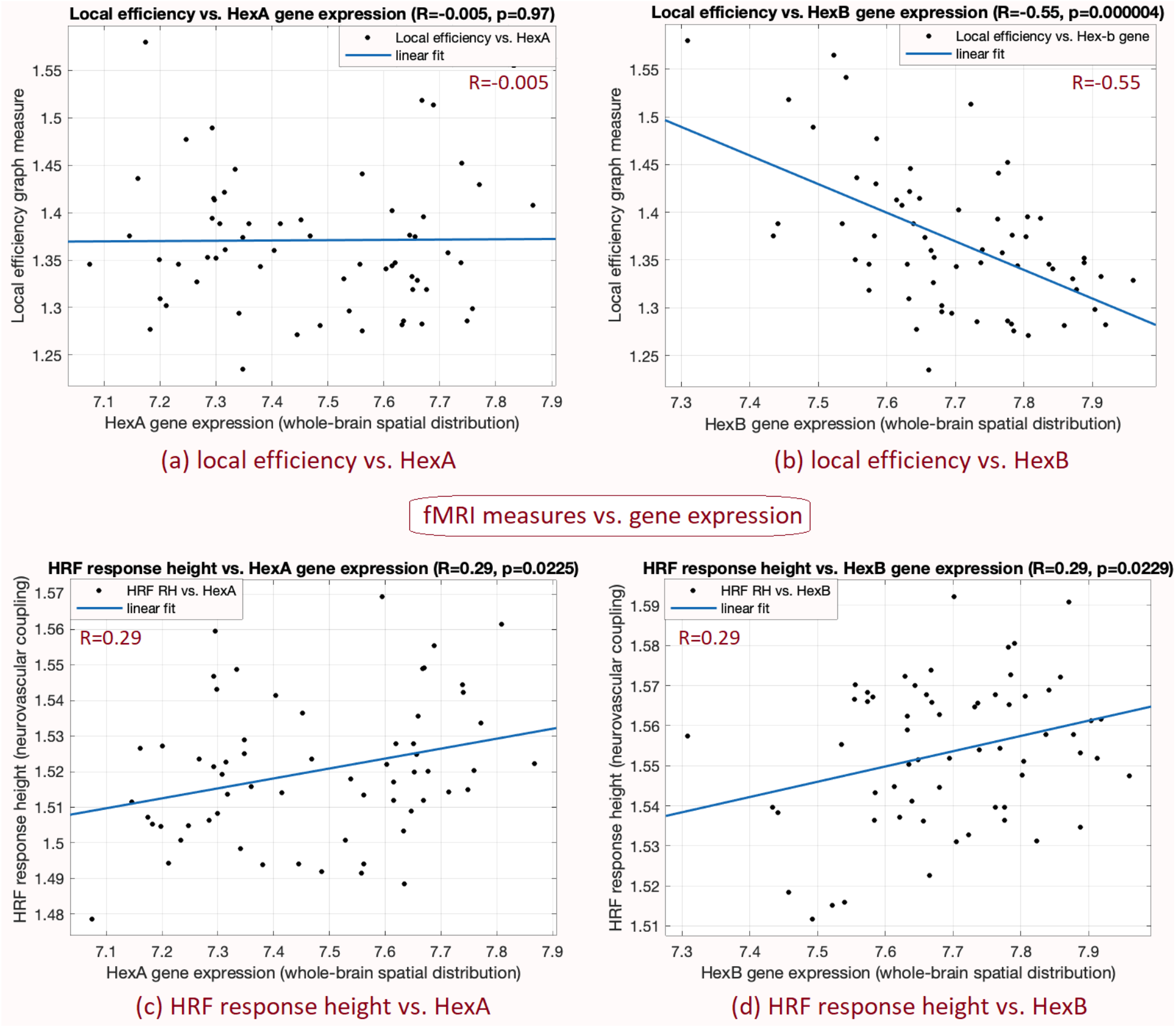
Associations between the spatial distribution of fMRI measures across the brain and the spatial distribution of gene expressions (HEXA in LOTS and HEXB in LOSD). (a) Local efficiency vs. HEXA in LOTS (R=−0.005, p=0.97), (b) local efficiency vs. HEXB in LOSD (R=−0.55, p=0.000004), (c) HRF response height vs. HEXA in LOTS (R=0.29, p=0.0225), (d) HRF response height vs. HEXB in LOSD (R=0.29, p=0.0229). Lower local efficiency and higher HRF RH were related to higher gene expression (with significant associations). Each data point corresponds to one of the 62 brain regions from the Desikan-Killiany whole-brain atlas.

**Figure 7.**
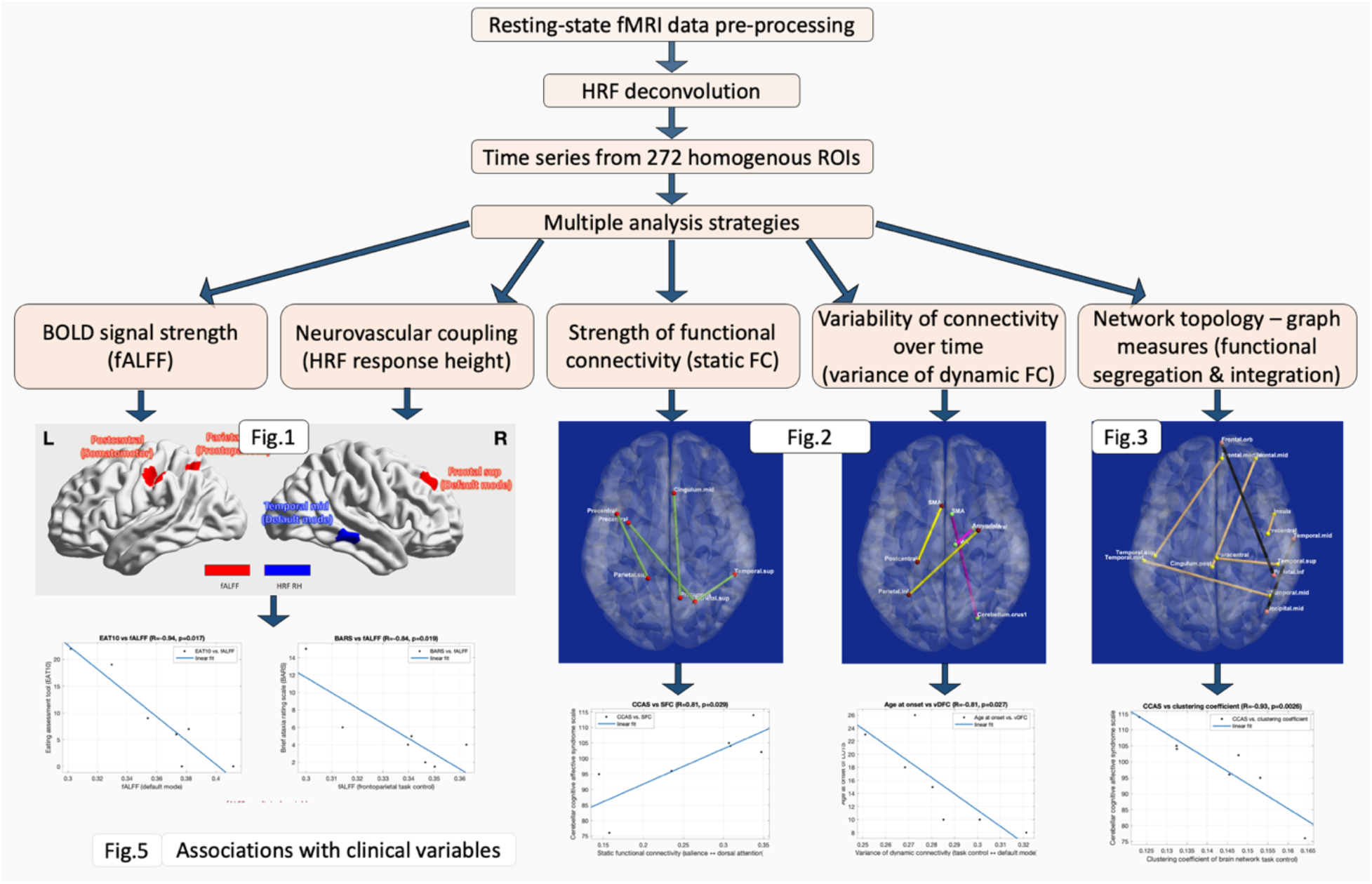
Flowchart illustrating our analysis strategy and corresponding results.

## 4. Discussion

As the first fMRI study of LOGG (or any other variant of GM2 gangliosidosis), we used a multi-dimensional approach by studying LOGG impairments in strength and variability of functional connectivity as well as brain network topology, BOLD signal strength, and neurovascular coupling. We found evidence for widespread cortical aberrations in LOGG and their relation to gene expression and clinical presentation. Amalgamating the different arms of our analysis framework, our findings converge to a single conclusion of cortical functional impairments in cognitive control, DMN, and somatomotor regions amongst the LOGG cohort. Cognitive control networks were featured in 4 of the 6 clinical associations, which taken together with imaging findings, underscore the importance of these networks (attention, task control, salience) for LOGG neurobiology. We posit that these may contribute to the psychiatric/cognitive symptoms presenting in a majority of LOGG patients.

Severe psychiatric presentations (such as psychosis and altered mood) have been reported in LOGG [13] [86] [20]. Significant cognitive impairment in memory, verbal, and executive function domains have also been reported [17] [14] [19] [20]. Deficits in processing speed, attention, and memory are also noted [18]. Oculomotor disturbances have also been observed [12], likely involving the dorsal attention network. Although cerebellar impairment has been the central observation in prior non-fMRI LOGG studies, extra-cerebellar abnormalities have also been described. These include cerebral white-matter hypo-density [21] and atrophy [31], reduced glucose metabolism in bilateral temporal and occipital lobes [22], and impaired occipital neuronal health [23]. With this background, we set out to discuss findings from each fMRI measure.

### FALFF

We found reduced fALFF in the DMN, somatomotor, and frontoparietal regions. Lower fALFF in the DMN has also been observed in other degenerative movement disorders, such as Parkinson’s disease [60], wherein it is interpreted as pathological malfunction of the region due to cellular breakdown. There has not been precise identification of the stated regions in the limited LOGG literature, but it is possible that ganglioside deposition could lead to cellular breakdown and subsequently reduced fALFF. This should be further investigated using multimodal imaging.

### HRF

Although impaired HRF has been noted in psychiatric conditions [87] [78] [51] [62] [88] [89] [90], this is, to the best of our knowledge, the first study to report HRF impairments in a neurological disease. We observed higher HRF RH in a middle temporal (DMN) region. We do not have a reference for HRF aberrations in similar neurological diseases. However, higher RH has been reported in psychiatric disorders such as posttraumatic stress disorder [91], attributed to disrupted metabolism arising from pathology [89]. It is possible that a similar mechanism in a metabolic disease like LOGG leads to impaired neurovascular coupling. In fact, one LOGG study has already reported reduced glucose metabolism in the temporal cortex [22].

### SFC

We observed pathologically higher SFC in LOGG, almost exclusively in cognitive control networks (mainly the dorsal attention network). SFC has been a robust marker of neurological disease pathology [1] [2] [3] [60], and such hyper-connectivity has been frequently reported in several diseases [56] [92]. The understanding is that fMRI hyper-connectivity is a result of neurological disruption [92]. Neuronal dysfunction in these regions and/or damage to connecting white-matter fibers results in aberrant communication between them – sometimes observed as reduced higher frequency electrical activity and increased lower frequency activity (local field potential from invasive electrodes) [93] [94]. The pathological low frequency activity in these regions presents as higher synchronization between them and thus higher fMRI connectivity [93] [94] [95]. In LOGG, both white-matter lesions [33] and lower white-matter density [21] have been reported in the cortex, coupled with cortical atrophy [31] and neuronal damage, possibly linked to ganglioside deposition [9]. Interestingly, the dorsal attention network was most affected by higher SFC. This network is chiefly responsible for visual attention and oculomotor function. Oculomotor disturbances have been previously described in LOGG but were not part of our current assessment [12]. Overall, our SFC result hints at a model of neuronal dysfunction (through ganglioside deposition and/or cortical atrophy) and/or white matter hypodensity/lesions, leading to pathologically higher connectivity in the dorsal attention network (and some other cognitive control networks), possibly resulting in oculomotor disturbances in LOGG. Additionally, since other cognitive control networks were also involved here, higher SFC might also contribute to the presentation of cognitive impairments and psychiatric presentations observed in almost half of all LOGG patients [13] [14] [17] [18]. Interestingly, SFC between salience and dorsal attention networks was significantly associated with CCAS (R=0.81), a measure of executive function, spatial cognition and affect regulation deficits. Pathologically higher SFC was related to worse symptoms. However, given our small sample size, it is possible that we missed the detection of some other important SFC impairments, which future studies in larger samples must probe further.

### DFC

Impairments in vDFC were observed in frontal and parietal regions associated largely with cognitive control networks. The supplementary motor area was notably involved in 3 of the 4 connections. DFC provides fundamentally different information from SFC [66] [65]. Psychiatric disorders largely present with reduced vDFC [56] [70] [72], although higher vDFC has been reported once [96]. DFC research has not been able to conclude the precise meaning of lower vDFC yet, but mounting evidence suggests that lower vDFC indicates the inability to dynamically adjust mental processes to changing body and environmental conditions [56] [70] [78] [97] [98] [99] [100]. The meaning of pathologically higher vDFC remains unknown. We found reduced vDFC of dorsal attention and somatomotor networks with cognitive control networks, which again might relate to oculomotor disturbances and cognitive impairments in LOGG. Lower vDFC is a biomarker of poorer cognitive performance [69] [70] [72] and worse psychiatric symptoms [56]. It remains unclear what higher vDFC seen in two other connections means, although one study has reported higher vDFC in a psychiatric condition before [96]. One such connection between supplementary motor area and cerebellum crus 1 was significantly associated with the age of disease onset (R=−0.81), with earlier onset related to pathologically higher vDFC between these motor-related regions.

### Graph measures

These measures have shown sensitivity to neurological disease pathology [1] [73] [74]. We found higher segregation in a prefrontal task control region, implying pathological over-connectivity between this region and the rest of the brain. We also found 7 connections exhibiting impaired functional integration with cognitive control networks dominating our fMRI findings. Summing all brain regions assessed from Tables 2 through 6, 21 of the 38 regions belonged to cognitive control networks. They also made up a third of the regions involved in impaired integration. Two additional important observations remain regarding the DMN (9 of 38 regions) and somatomotor network (6 of 38 regions). The DMN mainly exhibited impaired functional integration in LOGG (6 regions), although it did not feature in SFC or vDFC impairments. The DMN is involved in self-referential processing and plays a role in switching between task-positive and task-negative networks depending on external circumstances (task switching) [5] [95] [101]. Impaired DMN activity and connectivity have been linked to most psychiatric and neurological conditions [102] [103]. The fact that we did not find SFC/vDFC impairment in DMN but saw lower integration implies that specific DMN connections were not sufficiently affected (sub-threshold, not crossing our SFC/vDFC significance threshold). However, they had a significant impact on the remainder of the brain network through reduced functional integration. Both DMN-DMN and DMN-task control connections were impacted, implying that both self-referential processing and task switching were likely affected, respectively. In fact, the shortest path length between DMN and task control networks was significantly associated with disease duration (R=0.84), with a higher value (poorer integration) related to longer disease duration. The DMN also showed impaired fALFF and HRF. As in psychosis [104] and depression [105], these DMN impairments may contribute to the psychosis and mood disorders in LOGG [13] [86]. Unfortunately, we did not acquire validated clinical metrics of psychiatric disease from our patients to correlate with the functional results. Cognitive impairments in memory and processing speed also involve DMN aberrations [106] [107] and were also observed in LOGG [17] [14] [18] [19], which may also be explained by the DMN impairments. The somatosensory/motor network also had impaired connectivity and integration, which may relate to the dominant ataxia symptoms in LOGG. Although none of the ataxia scales correlated with somatomotor fMRI findings, future studies with larger sample sizes must not discount this possibility because ataxia is among the primary clinical presentations in LOGG, particularly in LOTS [9].

Substantial lateralization was noticed in graph measures, with 100% of segregation regions being in the left hemisphere and 79% of integration regions involving the right hemisphere. Additionally, a unanimous pattern of higher segregation and lower integration was observed, implying that LOGG is characterized by an imbalance in segregation/integration. The fine balance between segregation and integration (metastability) [108], is essential for healthy cognitive functioning [108], and is destabilized in psychiatric [74] [79] and neurological disorders [73]. In these disorders, the destabilized profile is that of higher segregation (pathologically elevated localized processing) and lower integration (pathologically reduced communication between localized subnetworks). This profile was also observed in LOGG.

### Cerebellum

The absence of cerebellar abnormalities in our results (except one vDFC connection) is notable. While structural and metabolic cerebellar abnormalities are a hallmark of LOGG [24] [26] [27] [22] [28] [29] [30], functional impairments observed in this study were dominated by cortical regions, mainly in cognitive control but also DMN and somatomotor regions. This implies damage to the brain beyond known structural or metabolic abnormalities, and affecting regions other than the cerebellum. Such damage could help explain the etiology of psychiatric, cognitive, and motor issues in this debilitating disease.

### Associations with gene expression

LOGG is a genetic disorder [12], and *HEXA* and *HEXB* gene expression were significantly associated with HRF RH in respective groups (R=0.29 and 0.29) and explained 8.5% of the variance in RH. Pathologically higher RH was related to higher gene expression. Interestingly, genes influence RH and TTP of the HRF (taken together) by 24– 51% [63] in healthy adults. Although we do not know more about this gene-HRF link in LOGG, it can be postulated that dysfunction in these genes may contribute to pathological amplitude of neurovascular coupling (which is, in turn, modulated by metabolites [109] [110]). Local efficiency was also associated with *HEXB* gene expression in LOSD (R=−0.55), with reduced ROI-brain communication related to higher gene expression.

The Allen brain atlas of gene expression [80] has opened new doors in imaging genetics. Our approach, involving correlating spatial maps of gene expression and imaging data, is gaining attention in recent years [83] [84]. Although using gene expression maps generated from a normative healthy sample and not our disease group is not the ideal approach, the alternative approach of obtaining genetic data from blood samples is also marred with imperfections as it does not inform us of gene expression in relevant brain regions (or even generally in the brain for that matter). Both approaches may be complementary, as one has spatial specificity and the other patient specificity. Thus, future studies in LOGG should probe gene-imaging relationship in greater detail.

### Limitations

Our study has three main limitations: (i) A small sample size. However, we have a relatively large cohort compared to prior literature on this disease (typically N=1–3). Sample sizes are commonly small because of the ultra-rare nature of LOGG (prevalence≈0.0003% [111]). Despite this, the current study is among the largest studies to have ever been conducted in LOGG with any imaging modality. (ii) We were unable to provide more granular differentiation between LOTS and LOSD as there were only two LOSD patients. (iii) Our data with TR=2s did not have satisfactory temporal resolution as HRF RH and DFC are particularly impacted by sampling rate. HRF TTP and FWHM were therefore ignored. The sensitivity of DFC to faster variations was also reduced [76] [77]. Despite these limitations, our findings provide a valuable starting point for future researchers to conduct more advanced hypothesis-driven studies in LOGG, ultimately for superior diagnostic assessments, disease monitoring, and therapeutic interventions.

## 5. Conclusions

In this first fMRI study of LOGG, we found widespread cortical aberrations chiefly in cognitive control networks (attention, task control), but also in the default mode and somatomotor networks. Some of these findings were significantly associated with certain clinical variables and gene expression of *HEXA* and *HEXB*. These functional aberrations might contribute to the psychiatric, cognitive, and oculomotor issues observed in the majority of LOGG patients. These primarily cerebral findings represent a paradigm shift from cerebellar structural and metabolic impairments reported in earlier LOGG studies. Future studies with larger samples are needed to assess functional impairments further and look for mechanistic insights beyond the cerebellum.

## Data Availability

Late-onset GM2 gangliosidosis (LOGG) is an extremely rare disease. Hence, the detailed protocol (approved by the Mass General Brigham IRB) states that all de-identifiable imaging and clinical data and any other study data can be shared externally only with both IRB approval and a data usage agreement.

## CRediT (Contributor Roles Taxonomy) – author contributions

**D Rangaprakash**: Conceptualization, Methodology, Software, Data Analysis, Investigation, Visualization, Writing - Original Draft, Reviewing and Editing. **Olivia E Rowe**: Data Acquisition, Investigation, Writing - Reviewing and Editing. **Christopher D Stephen**: Data Acquisition, Investigation, Writing - Reviewing and Editing. **Florian S Eichler**: Funding Acquisition, Data Acquisition, Investigation, Writing - Reviewing and Editing. **Eva-Maria Ratai**: Funding Acquisition, Project Administration, Investigation, Writing - Reviewing and Editing, Supervision. **Robert L Barry**: Funding Acquisition, Resources, Project Administration, Conceptualization, Methodology, Data Acquisition, Investigation, Writing - Reviewing and Editing, Supervision.

## Acknowledgments

We thank the patients (and their families) for their participation in this research. This research was supported by National Tay-Sachs & Allied Diseases Association Inc., and the Sanofi US Services Inc. MRI was performed at the Athinoula A. Martinos Center for Biomedical Imaging using resources provided by the Center for Mesoscale Mapping (P41EB030006) and the Center for Functional Neuroimaging Technologies (P41EB015896), as well as Biotechnology Resource Grants supported by the National Institute of Biomedical Imaging and Bioengineering, and the National Institutes of Health (NIH). The NIH also extended support through grants R00EB016689 and R01EB027779 (to R.L.B.). This study was also supported in part by the Athinoula A. Martinos Center for Biomedical Imaging. This content is solely the responsibility of the authors and does not necessarily reflect the official views of the NIH.

## Data Availability Statement

LOGG is an extremely rare disease. Hence, the detailed protocol (approved by the Mass General Brigham IRB) states that all de-identifiable imaging and clinical data and any other study data can be shared externally only with both IRB approval and a data usage agreement.

## References

[1] S. Zhao, D. Rangaprakash, P. Liang and G. Deshpande, “Deterioration from healthy to mild cognitive impairment and Alzheimer’s disease mirrored in corresponding loss of centrality in directed brain networks,” Brain Informatics, vol. 6, no. 1, p. 8, 2019.

[2] L. Solstrand Dahlberg, O. Lungu and J. Doyon, “Cerebellar Contribution to Motor and Non-motor Functions in Parkinson’s Disease: A Meta-Analysis of fMRI Findings,” Front Neurol., vol. 11, p. 127, 2020.

[3] B. Conrad, R. Barry, B. Rogers, S. Maki, A. Mishra, S. Thukral, S. Sriram, A. Bhatia, S. Pawate, J. Gore and S. Smith, “Multiple sclerosis lesions affect intrinsic functional connectivity of the spinal cord,” Brain, vol. 141, no. 6, pp. 1650–1664, 2018.

[4] J. Belliveau, D. Kennedy, R. McKinstry, B. Buchbinder, R. Weisskoff, M. Cohen, J. Vevea, T. Brady and B. Rosen, “Functional mapping of the human visual cortex by magnetic resonance imaging,” Science, vol. 254, no. 5032, pp. 716–9, 1991.

[5] J. Power, A. Cohen, S. Nelson, G. Wig, K. Barnes, J. Church, A. Vogel, T. Laumann, F. Miezin, B. Schlaggar and S. Petersen, “Functional network organization of the human brain,” Neuron, vol. 72, no. 4, pp. 665–78, 2011.

[6] M. Rubinov and O. Sporns, “Complex network measures of brain connectivity: uses and interpretations,” Neuroimage, vol. 52, no. 3, pp. 1059–69, 2010.

[7] R. Tadayonnejad, S. Yang, A. Kumar and O. Ajilore, “Clinical, cognitive, and functional connectivity correlations of resting-state intrinsic brain activity alterations in unmedicated depression,” Journal of Affective Disorders, vol. 172, pp. 241–50, 2015.

[8] D. Rangaprakash, M. N. Dretsch, W. Yan, J. S. Katz, T. S. Denney and G. Deshpande, “Hemodynamic variability in soldiers with trauma: Implications for functional MRI connectivity studies,” NeuroImage: Clinical, vol. 16, pp. 409–417, 2017.

[9] A. Percy, L. Shapiro and M. Kaback, “Inherited lipid storage diseases of the central nervous system,” Curr Probl Pediatr., vol. 9, no. 11, pp. 1–51, 1979.

[10] C. Toro, M. Zainab and C. Tifft, “The GM2 gangliosidoses: Unlocking the mysteries of pathogenesis and treatment.,” Neuroscience letters, vol. 764, p. 136195, 2021.

[11] “Tay Sachs Disease,” NORD (National Organization for Rare Disorders), 2017. [Online]. Available: https://rarediseases.org/rare-diseases/tay-sachs-disease/.

[12] C. Stephen, D. Balkwill, P. James, E. Haxton, K. Sassower, J. Schmahmann, F. Eichler and R. Lewis, “Quantitative oculomotor and nonmotor assessments in late-onset GM2 gangliosidosis,” Neurology, vol. 94, no. 7, pp. e705–e717, 2020.

[13] M. Masingue, L. Dufour, T. Lenglet, L. Saleille, C. Goizet, X. Ayrignac, F. Ory-Magne, M. Barth, F. Lamari, D. Mandia, C. Caillaud and Y. Nadjar, “Natural History of Adult Patients with GM2 Gangliosidosis,” Ann Neurol., vol. 87, no. 4, pp. 609–617, 2020.

[14] L. Frey, S. Ringel and C. Filley, “The natural history of cognitive dysfunction in late-onset GM2 gangliosidosis,” Arch Neurol., vol. 62, no. 6, pp. 989–94, 2005.

[15] D. Hamilton, “A nursing challenge: adult-onset Tay-Sachs disease,” Arch Psychiatr Nurs., vol. 5, no. 6, pp. 382–5, 1991.

[16] B. Shapiro, S. Hatters-Friedman, J. Fernandes-Filho, K. Anthony and M. Natowicz, “Late-onset Tay-Sachs disease: adverse effects of medications and implications for treatment,” Neurology, vol. 67, no. 5, pp. 875–7, 2006.

[17] C. Zaroff, O. Neudorfer, C. Morrison, G. Pastores, H. Rubin and E. Kolodny, “Neuropsychological assessment of patients with late onset GM2 gangliosidosis,” Neurology, vol. 62, no. 12, pp. 2283–6, 2004.

[18] A. Barritt, S. Anderson, P. Leigh and B. Ridha, “Late-onset Tay-Sachs disease,” Pract Neurol., vol. 17, no. 5, pp. 396–399, 2017.

[19] D. Elstein, G. Doniger, E. Simon, I. Korn-Lubetzki, R. Navon and A. Zimran, “Neurocognitive testing in late-onset Tay-Sachs disease: a pilot study,” J Inherit Metab Dis., vol. 31, no. 4, pp. 518–23, 2008.

[20] G. MacQueen, P. Rosebush and M. Mazurek, “Neuropsychiatric aspects of the adult variant of Tay-Sachs disease,” J Neuropsychiatry Clin Neurosci., vol. 10, no. 1, pp. 10–19, 1998.

[21] M. Fukumizu, H. Yoshikawa, S. Takashima, N. Sakuragawa and T. Kurokawa, “Tay-Sachs disease: progression of changes on neuroimaging in four cases,” Neuroradiology, vol. 34, no. 6, pp. 483–6, 1992.

[22] Z. Jamrozik, A. Lugowska, M. Gołębiowski, L. Królicki, J. Mączewska and M. KuŹma-Kozakiewicz, “Late onset GM2 gangliosidosis mimicking spinal muscular atrophy,” Gene, vol. 527, no. 2, pp. 679–82, 2013.

[23] M. Inglese, A. Nusbaum, G. Pastores, J. Gianutsos, E. Kolodny and O. Gonen, “MR imaging and proton spectroscopy of neuronal injury in late-onset GM2 gangliosidosis,” AJNR Am J Neuroradiol., vol. 26, no. 8, pp. 2037–42, 2005.

[24] O. E. Rowe, D. Rangaprakash, A. Weerasekera, N. Godbole, E. Haxton, P. F. James, C. D. Stephen, R. L. Barry, F. S. Eichler and E. M. Ratai, “Magnetic resonance imaging and spectroscopy in late-onset GM2-gangliosidosis.,” Molecular genetics and metabolism, vol. 133, no. 4, p. 386–396, 2021.

[25] S. Grosso, M. Farnetani, R. Berardi, M. Margollicci, P. Galluzzi, R. Vivarelli, G. Morgese and P. Ballestri, “GM2 gangliosidosis variant B1 neuroradiological findings,” J Neurol., vol. 250, no. 1, pp. 17–21, 2003.

[26] A. Deik and R. Saunders-Pullman, “Atypical presentation of late-onset Tay-Sachs disease,” Muscle Nerve., vol. 49, no. 5, pp. 768–71, 2014.

[27] O. Neudorfer, G. Pastores, B. Zeng, J. Gianutsos, C. Zaroff and E. Kolodny, “Late-onset Tay-Sachs disease: phenotypic characterization and genotypic correlations in 21 affected patients,” Genet Med., vol. 7, no. 2, pp. 119–23, 2005.

[28] H. Jahnová, H. Poupětová, J. Jirečková, H. Vlášková, E. Košťálová, R. Mazanec, A. Zumrová, P. Mečíř, Z. Mušová and M. Magner, “Amyotrophy, cerebellar impairment and psychiatric disease are the main symptoms in a cohort of 14 Czech patients with the late-onset form of Tay-Sachs disease,” J Neurol., vol. 266, no. 8, pp. 1953–1959, 2019.

[29] J. Streifler, M. Gornish, H. Hadar and N. Gadoth, “Brain imaging in late-onset GM2 gangliosidosis,” Neurology, vol. 43, no. 10, pp. 2055–8, 1993.

[30] E. Hund, A. Grau, W. Fogel, M. Forsting, M. Cantz, B. Kustermann-Kuhn, K. Harzer, R. Navon, H. Goebel and H. Meinck, “Progressive cerebellar ataxia, proximal neurogenic weakness and ocular motor disturbances: hexosaminidase A deficiency with late clinical onset in four siblings,” J Neurol Sci., vol. 145, no. 1, pp. 25–31, 1997.

[31] K. Grim, G. Phillips and D. Renner, “Dysarthria and Stutter as Presenting Symptoms of Late-Onset Tay-Sachs Disease in Three Siblings.,” Movement disorders clinical practice, vol. 2, no. 3, p. 289–290, 2015.

[32] I. Prihodova, T. Kalincik, H. Poupetova, H. Jahnova and S. Nevsimalova, “Late-onset tay-sachs disease can mimic spinal muscular atrophy type III - Two case reports.,” Ces. a Slov. Neurol. a Neurochir., vol. 76, p. 221–224, 2013.

[33] C. Delnooz, D. Lefeber, S. Langemeijer, S. Hoffjan, G. Dekomien, M. Zwarts, B. Van Engelen, R. Wevers, H. Schelhaas and B. van de Warrenburg, “New cases of adult-onset Sandhoff disease with a cerebellar or lower motor neuron phenotype.,” Journal of neurology, neurosurgery, and psychiatry, vol. 81, no. 9, p. 968–972, 2010.

[34] J. Schmahmann, R. Gardner, J. MacMore and M. Vangel, “Development of a brief ataxia rating scale (BARS) based on a modified form of the ICARS.,” Movement disorders., vol. 24, no. 12, p. 1820–1828, 2009.

[35] T. Schmitz-Hübsch, S. du Montcel, L. Baliko, J. Berciano, S. Boesch, C. Depondt, P. Giunti, C. Globas, J. Infante, J. Kang, B. Kremer, C. Mariotti, B. Melegh, M. Pandolfo, M. Rakowicz, P. Ribai, R. Rola, L. Schöls and e. al., “Scale for the assessment and rating of ataxia: development of a new clinical scale.,” Neurology, vol. 66, no. 11, p. 1717–1720, 2006.

[36] S. Subramony, W. May, D. Lynch, C. Gomez, K. Fischbeck, M. Hallett, P. Taylor, R. Wilson, T. Ashizawa and C. A. Group, “Measuring Friedreich ataxia: Interrater reliability of a neurologic rating scale.,” Neurology, vol. 64, no. 7, p. 1261–1262, 2005.

[37] C. Toro, L. Shirvan and C. Tifft, “HEXA Disorders,” in Adam, M.P.; Ardinger, H.H.; Pagon, R.A.; Wallace, S.E.; Bean, L.J.H.; Stephens, K.; Amemiya, A.; editors; GeneReviews, Seattle, University of Washington, Seattle, 1999.

[38] F. Hoche, X. Guell, J. Sherman, M. Vangel and J. Schmahmann, “Cerebellar Contribution to Social Cognition.,” Cerebellum, vol. 15, no. 6, p. 732–743, 2016.

[39] F. Hoche, X. Guell, M. Vangel, J. Sherman and J. Schmahmann, “The cerebellar cognitive affective/Schmahmann syndrome scale.,” Brain, vol. 141, no. 1, p. 248–270, 2018.

[40] T. Mollayeva, P. Thurairajah, K. Burton, S. Mollayeva, C. M. Shapiro and A. Colantonio, “The Pittsburgh sleep quality index as a screening tool for sleep dysfunction in clinical and non-clinical samples: A systematic review and meta-analysis.,” Sleep medicine reviews, vol. 25, p. 52–73, 2016.

[41] M. Johns, “A new method for measuring daytime sleepiness: the Epworth sleepiness scale,” Sleep., vol. 14, no. 6, pp. 540–5, 1991.

[42] P. Belafsky, D. Mouadeb, C. Rees, J. Pryor, G. Postma, J. Allen and R. Leonard, “Validity and reliability of the Eating Assessment Tool (EAT-10).,” The Annals of otology, rhinology, and laryngology, vol. 117, no. 12, p. 919–924, 2008.

[43] S. Whitfield-Gabrieli and A. Nieto-Castanon, “Conn: A functional connectivity toolbox for correlated and anticorrelated brain networks,” Brain Connectivity, vol. 2, no. 3, pp. 125–141, 2012.

[44] K. J. Friston, J. Ashburner, S. J. Kiebel, T. E. Nichols and W. D. Penny, Statistical Parametric Mapping: The Analysis of Functional Brain Images, Academic Press, 2007.

[45] S. Whitfield-Gabrieli, “Artifact Detection Tools (ART),” 30 04 2008. [Online]. Available: https://www.nitrc.org/projects/artifact_detect. [Accessed 27 03 2020].

[46] J. Power, K. Barnes, A. Snyder, B. Schlaggar and S. Petersen, “Spurious but systematic correlations in functional connectivity MRI networks arise from subject motion,” Neuroimage, vol. 59, no. 3, pp. 2142–54, 2012.

[47] D. A. Handwerker, J. M. Ollinger and M. D’Esposito, “Variation of BOLD hemodynamic responses across subjects and brain regions and their effects on statistical analyses,” Neuroimage, vol. 21, no. 4, pp. 1639–51, 2004.

[48] D. Rangaprakash, G.-R. Wu, D. Marinazzo, X. Hu and G. Deshpande, “Hemodynamic response function (HRF) variability confounds resting-state fMRI functional connectivity,” Magnetic Resonance in Medicine, p. in press, 2018.

[49] D. Rangaprakash, G.-R. Wu, D. Marinazzo, X. Hu and G. Deshpande, “Hemodynamic Response Function Parameters Derived from Resting-State Functional MRI Data of Healthy Individuals Obtained in a 7T MRI Scanner,” Data in Brief, p. in press, 2018.

[50] M. E. Archila-Meléndez, C. Sorg and C. Preibisch, “Modeling the impact of neurovascular coupling impairments on BOLD-based functional connectivity at rest.,” NeuroImage, vol. 218, p. 116871, 2020.

[51] W. Yan, D. Rangaprakash and G. Deshpande, “Aberrant Hemodynamic Responses in Autism: Implications for Resting State fMRI Functional Connectivity Studies,” Neuroimage: Clinical, vol. 19, pp. 320–330, 2018.

[52] G. Wu, W. Liao, S. Stramaglia, J. Ding, H. Chen and D. Marinazzo, “A blind deconvolution approach to recover effective connectivity brain networks from resting state fMRI data,” Med Image Anal., vol. 17, no. 3, pp. 365–374, 2013.

[53] M. Boly, S. Sasai, O. Gosseries, M. Oizumi, A. Casali, M. Massimini and G. Tononi, “Stimulus set meaningfulness and neurophysiological differentiation: a functional magnetic resonance imaging study,” PLoS One, vol. 10, no. 5, p. e0125337, 2015.

[54] E. Amico, F. Gomez, C. Di Perri, A. Vanhaudenhuyse, D. Lesenfants, P. Boveroux, V. Bonhomme, J. F. Brichant, D. Marinazzo and S. Laureys, “Posterior cingulate cortex-related co-activation patterns: a resting state FMRI study in propofol-induced loss of consciousness,” PLoS One, vol. 9, no. 6, p. e100012, 2014.

[55] D. Rangaprakash, M. N. Dretsch, W. Yan, J. S. Katz, T. S. Denney and G. Deshpande, “Hemodynamic response function parameters obtained from resting-state functional MRI data in soldiers with trauma,” Data in Brief, vol. 14, pp. 558–562, 2017.

[56] D. Rangaprakash, G. Deshpande, T. Daniel, A. Goodman, J. Robinson, N. Salibi, J. Katz, T. Denney and M. Dretsch, “Compromised Hippocampus-Striatum Pathway as a Potential Imaging Biomarker of Mild Traumatic Brain Injury and Posttraumatic Stress Disorder,” Human Brain Mapping, vol. 38, no. 6, pp. 2843–2864, 2017.

[57] B. Lamichhane, B. M. Adhikari, S. F. Brosnan and M. Dhamala, “The neural basis of perceived unfairness in economic exchanges,” Brain Connectivity, vol. 4, no. 8, pp. 619–30, 2014.

[58] J. Frazier, S. Chiu, J. Breeze, N. Makris, N. Lange, D. Kennedy, M. Herbert, E. Bent, V. Koneru, M. Dieterich, S. Hodge, S. Rauch, P. Grant, B. Cohen, L. Seidman, V. Caviness and J. Biederman, “Structural brain magnetic resonance imaging of limbic and thalamic volumes in pediatric bipolar disorder,” Am J Psychiatry, vol. 162, no. 7, pp. 1256–65, 2005.

[59] R. Buckner, F. Krienen, A. Castellanos, J. Diaz and B. Yeo, “The organization of the human cerebellum estimated by intrinsic functional connectivity,” J Neurophysiol., vol. 106, no. 5, pp. 2322–45, 2011.

[60] R. Franciotti, S. Delli Pizzi, M. Russo, C. Carrarini, D. Carrozzino, B. Perfetti, M. Onofrj and L. Bonanni, “Somatic symptoms disorders in Parkinson’s disease are related to default mode and salience network dysfunction,” Neuroimage Clin., vol. 23, p. 101932, 2019.

[61] X. Du, J. Liu, Q. Hua and Y. Wu, “Relapsing-Remitting Multiple Sclerosis Is Associated With Regional Brain Activity Deficits in Motor- and Cognitive-Related Brain Areas,” Front Neurol., vol. 10, p. 1136, 2019.

[62] D. Rangaprakash, R. Tadayonnejad, G. Deshpande, J. O’Neill and J. Feusner, “FMRI hemodynamic response function (HRF) as a novel marker of brain function: applications for understanding obsessive-compulsive disorder pathology and treatment response,” Brain Imaging and Behavior, p. in press, 2020.

[63] Z. Shan, A. Vinkhuyzen, P. Thompson, K. McMahon, G. Blokland, G. de Zubicaray, V. Calhoun, N. Martin, P. Visscher, M. Wright and D. Reutens, “Genes influence the amplitude and timing of brain hemodynamic responses,” NeuroImage, vol. 124, no. Pt A, pp. 663–671, 2016.

[64] P. Lin, J. Sun, G. Yu, Y. Wu, Y. Yang, M. Liang and X. Liu, “Global and local brain network reorganization in attention-deficit/hyperactivity disorder,” Brain Imaging and Behavior, vol. 8, no. 4, pp. 558–69, 2014.

[65] R. Hutchison, T. Womelsdorf, E. Allen, P. Bandettini, V. Calhoun, M. Corbetta, S. Della-Penna, J. Duyn, G. Glover, J. Gonzalez-Castillo, D. Handwerker, S. Keilholz, V. Kiviniemi, D. Leopold, F. de Pasquale, O. Sporns, M. Walter and C. Chang, “Dynamic functional connectivity: promise, issues, and interpretations.,” Neuroimage, vol. 80, pp. 360–378, 2013.

[66] E. Hansen, D. Battaglia, A. Spiegler, G. Deco and V. Jirsa, “Functional connectivity dynamics: modeling the switching behavior of the resting state,” Neuroimage, vol. 105, pp. 525–35, 2015.

[67] W. Majeed, M. Magnuson, W. Hasenkamp, H. Schwarb, E. Schumacher, L. Barsalou and S. Keilholz, “Spatiotemporal dynamics of low frequency BOLD fluctuations in rats and humans,” NeuroImage, vol. 54, pp. 1140–1150, 2011.

[68] S. Keilholz, M. Magnuson, W. Pan., M. Willis and G. Thompson, “Dynamic properties of functional connectivity in the rodent.,” Brain Connectivity, vol. 3, no. 1, pp. 31–40, 2013.

[69] G. Thompson, M. Magnuson, M. Merritt, H. Schwarb, W. Pan, A. McKinley, L. Tripp, E. Schumacher and S. Keilholz, “Short-time windows of correlation between large-scale functional brain networks predict vigilance intraindividually and interindividually.,” Human Brain Mapping (in press), vol. 34, no. 12, pp. 3280–98, 2013.

[70] H. Jia, X. Hu and G. Deshpande, “Behavioral relevance of the dynamics of the functional brain connectome,” Brain Connectivity, vol. 4, no. 9, pp. 741–59, 2014.

[71] C. Jin, H. Jia, P. Lanka, D. Rangaprakash, L. Li, T. Liu, X. Hu and G. Deshpande, “Dynamic brain connectivity is a better predictor of PTSD than static connectivity,” Hum Brain Mapp., vol. 38, no. 9, pp. 4479–4496, 2017.

[72] U. Sakoğlu, G. Pearlson, K. Kiehl, Y. Wang, A. Michael and V. Calhoun, “A method for evaluating dynamic functional network connectivity and task-modulation: application to schizophrenia.,” MAGMA, vol. 23, no. 5-6, pp. 351–366, 2010.

[73] M. A. Rocca, P. Valsasina, A. Meani, A. Falini, G. Comi and M. Filippi, “Impaired functional integration in multiple sclerosis: a graph theory study,” Brain Structure and Function, p. (in press), Sept 2014.

[74] Q. Yu, J. Sui, K. A. Kiehl, G. Pearlson and V. D. Calhoun, “State-related functional integration and functional segregation brain networks in schizophrenia,” Schizophrenia Research, vol. 150, no. 2–3, pp. 450–8, 2013.

[75] Q. Zou, C. Zhu, Y. Yang, X. Zuo, X. Long, Q. Cao, Y. Wang and Y. Zang, “An improved approach to detection of amplitude of low-frequency fluctuation (ALFF) for resting-state fMRI: fractional ALFF.,” J Neurosci Methods, vol. 172, no. 1, pp. 137–41, 2008.

[76] R. Hutchison, T. Womelsdorf, E. Allen, P. Bandettini, V. Calhoun, M. Corbetta, P. S. Della, J. Duyn, G. Glover, J. Gonzalez-Castillo and e. al., “Dynamic functional connectivity: promise, issues, and interpretations.,” Neuroimage, vol. 80, pp. 360–378, 2013.

[77] T. Shi, D. Rangaprakash and G. Deshpande, “Assessing the Reliability of Estimated Correlation during the Evaluation of Dynamic Functional Connectivity,” in Proceedings of the Annual Meeting of the International Society for Magnetic Resonance in Medicine (ISMRM), Singapore, 2016.

[78] D. Rangaprakash, M. Dretsch, A. Venkatraman, J. Katz, T. Denney and G. Deshpande, “Identifying Disease Foci from Static and Dynamic Effective Connectivity Networks: Illustration in Soldiers with Trauma,” Human Brain Mapping, vol. 39, no. 1, pp. 264–287, 2018.

[79] D. Rangaprakash, M. Dretsch, J. Katz, T. Denney and G. Deshpande, “Dynamics of Segregation and Integration in Directional Brain Networks: Illustration in Soldiers With PTSD and Neurotrauma.,” Front Neurosci., vol. 13, p. 803, 2019.

[80] M. Hawrylycz, E. Lein, A. Guillozet-Bongaarts, E. Shen, L. Ng and e. al., “An anatomically comprehensive atlas of the adult human brain transcriptome,” Nature, vol. 489, no. 7416, pp. 391–399, 2012.

[81] L. French and T. Paus, “A FreeSurfer view of the cortical transcriptome generated from the Allen Human Brain Atlas,” Front Neurosci., vol. 9, p. 323, 2015.

[82] R. Desikan, F. Ségonne, B. Fischl, B. Quinn, B. Dickerson, D. Blacker, R. Buckner, A. Dale, R. Maguire, B. Hyman, M. Albert and R. Killiany, “An automated labeling system for subdividing the human cerebral cortex on MRI scans into gyral based regions of interest,” NeuroImage, vol. 31, no. 3, pp. 968–80, 2006.

[83] M. McCarthy, S. Liang, A. Spadoni, J. Kelsoe and A. Simmons, “Whole brain expression of bipolar disorder associated genes: structural and genetic analyses.,” PLoS one, vol. 9, no. 6, p. e100204, 2014.

[84] G. Roshchupkin, H. Adams, S. van der Lee, M. Vernooij, C. van Duijn, A. Uitterlinden, A. van der Lugt, A. Hofman, W. Niessen and M. Ikram, “Fine-mapping the effects of Alzheimer’s disease risk loci on brain morphology.,” Neurobiology of aging, vol. 48, pp. 204–211, 2016.

[85] M. Xia, J. Wang and Y. He, “BrainNet Viewer: A Network Visualization Tool for Human Brain Connectomics,” PLoS One, vol. 8, p. e68910, 2013.

[86] J. Streifler, M. Golomb and N. Gadoth, “Psychiatric features of adult GM2 gangliosidosis,” Br J Psychiatry., vol. 155, pp. 410–413, 1989.

[87] D. Rangaprakash, M. N. Dretsch, W. Yan, J. S. Katz, T. S. Denney and G. Deshpande, “Hemodynamic variability in soldiers with trauma: Implications for functional MRI connectivity studies,” NeuroImage: Clinical, vol. 16, pp. 409–417, 2017.

[88] G. Deshpande, D. Rangaprakash, W. Yan, P. Liddle and L. Palaniyappan, “Characterization of Hemodynamic Alterations in Schizophrenia and Bipolar Disorder and their Effect on Resting-state Functional Connectivity,” in Schizophrenia International Research Society Conference (SIRS), Florence, Italy, April 2018.

[89] A. R. Mayer, T. Toulouse, S. Klimaj, J. M. Ling, A. Pena and P. S. Bellgowan, “Investigating the properties of the hemodynamic response function after mild traumatic brain injury.,” Journal of neurotrauma, vol. 31, no. 2, p. 189–197, 2014.

[90] I. G. Elbau, B. Brücklmeier, M. Uhr, J. Arloth, D. Czamara, V. I. Spoormaker, M. Czisch, K. E. Stephan, E. B. Binder and P. G. Sämann, “The brain’s hemodynamic response function rapidly changes under acute psychosocial stress in association with genetic and endocrine stress response markers.,” Proceedings of the National Academy of Sciences of the United States of America, vol. 115, no. 43, p. E10206–E10215, 2018.

[91] D. Rangaprakash, M. Dretsch, W. Yan, J. Katz, T. Denney and G. Deshpande, “Hemodynamic variability in soldiers with trauma: Implications for functional MRI connectivity studies,” NeuroImage: Clinical, vol. 16, pp. 409–417, 2017.

[92] F. Hillary, C. Roman, U. Venkatesan and et.al, “Hyperconnectivity is a fundamental response to neurological disruption,” Neuropsychology, vol. 29, no. 1, pp. 59–75, 2015.

[93] C. Canella, F. Rocchi, S. Noei, D. Gutierrez-Barragan, L. Coletta, A. Galbusera, S. Vassanelli, M. Pasqualetti, G. Iurilli, S. Panzeri and A. Gozzi, “Cortical silencing results in paradoxical fMRI overconnectivity.,” bioRxiv, vol. 237958, p. DOI: 10.1101/2020.08.05.237958, 2020.

[94] P. Drew, C. Mateo, K. Turner, X. Yu and D. Kleinfeld, “Ultra-slow Oscillations in fMRI and Resting-State Connectivity: Neuronal and Vascular Contributions and Technical Confounds.,” Neuron, vol. 107, no. 5, p. 782–804, 2020.

[95] H. Laufs, K. Krakow, P. Sterzer, E. Eger, A. Beyerle, A. Salek-Haddadi and A. Kleinschmidt, “Electroencephalographic signatures of attentional and cognitive default modes in spontaneous brain activity fluctuations at rest.,” Proceedings of the National Academy of Sciences of the United States of America, vol. 100, no. 19, p. 11053–11058, 2003.

[96] H. Bednarz, D. Rangaprakash, G. Deshpande and R. Kana, “Increased Intra-Individual Neural Variability in Autism.,” Archives of Clinical Neuropsychology, Vols. 31(B-19), pp. 584–673, 2016.

[97] L. Douw, D. G. Wakeman, N. Tanaka, H. Liu and S. M. Stufflebeam, “State-dependent variability of dynamic functional connectivity between frontoparietal and default networks relates to cognitive flexibility.,” Neuroscience, vol. 339, p. 12–21, 2016.

[98] B. Rashid, M. Arbabshirani, E. Damaraju, M. Cetin, R. Miller, G. Pearlson and V. Calhoun, “Classification of schizophrenia and bipolar patients using static and dynamic resting-state fMRI brain connectivity,” NeuroImage, vol. 134, pp. 645–657, 2016.

[99] D. Garrett, G. Samanez-Larkin, S. MacDonald, U. Lindenberger, A. McIntosh and C. Grady, “Moment-to-moment brain signal variability: a next frontier in human brain mapping?,” Neuroscience and Biobehavioral Reviews, vol. 37, no. 4, pp. 610–24, 2013.

[100] L. Li, Y. Wang, L. Ye, W. Chen, X. Huang, Q. Cui, Z. He, D. Liu and H. Chen, “Altered Brain Signal Variability in Patients With Generalized Anxiety Disorder.,” Frontiers in psychiatry, vol. 10, p. 84, 2019.

[101] L. Mak, L. Minuzzi, G. MacQueen, G. Hall, S. Kennedy and R. Milev, “The Default Mode Network in Healthy Individuals: A Systematic Review and Meta-Analysis.,” Brain connectivity, vol. 7, no. 1, p. 25–33, 2017.

[102] S. Whitfield-Gabrieli and J. Ford, “Default mode network activity and connectivity in psychopathology.,” Annual review of clinical psychology, vol. 8, p. 49–76, 2012.

[103] A. Mohan, A. Roberto, A. Mohan, A. Lorenzo, K. Jones, M. Carney, L. Liogier-Weyback, S. Hwang and K. Lapidus, “The Significance of the Default Mode Network (DMN) in Neurological and Neuropsychiatric Disorders: A Review.,” The Yale journal of biology and medicine, vol. 89, no. 1, p. 49–57, 2016.

[104] D. Wotruba, L. Michels, R. Buechler, S. Metzler, A. Theodoridou, M. Gerstenberg, S. Walitza, S. Kollias, W. Rössler and K. Heekeren, “Aberrant coupling within and across the default mode, task-positive, and salience network in subjects at risk for psychosis.,” Schizophrenia bulletin, vol. 40, no. 5, p. 1095–1104, 2014.

[105] H. Zhou, X. Chen, Y. Shen, L. Li, N. Chen, Z. Zhu, F. Castellanos and C. Yan, “Rumination and the default mode network: Meta-analysis of brain imaging studies and implications for depression.,” NeuroImage, vol. 206, p. 116287, 2020.

[106] J. Smallwood, B. Bernhardt, R. Leech, D. Bzdok, E. Jefferies and D. Margulies, “The default mode network in cognition: a topographical perspective.,” Nature reviews. Neuroscience, vol. 22, no. 8, p. 503–513, 2021.

[107] K. Mevel, B. Grassiot, G. Chételat, G. Defer, B. Desgranges and F. Eustache, “The default mode network: cognitive role and pathological disturbances,” Revue neurologique, vol. 166, no. 11, p. 859–872, 2010.

[108] P. Hellyer, G. Scott, M. Shanahan, D. Sharp and R. Leech, “Cognitive Flexibility through Metastable Neural Dynamics Is Disrupted by Damage to the Structural Connectome,” The Journal of Neuroscience, vol. 35, no. 24, pp. 9050–63, 2015.

[109] S. D. Muthukumaraswamy, C. J. Evans, R. A. Edden, R. G. Wise and K. D. Singh, “Individual variability in the shape and amplitude of the BOLD-HRF correlates with endogenous GABAergic inhibition,” Human Brain Mapping, vol. 33, no. 2, pp. 455–65, 2012.

[110] Z. Cohen, G. Bonvento, P. Lacombe and E. Hamel, “Serotonin in the regulation of brain microcirculation,” Progress in Neurobiology, vol. 50, no. 4, pp. 335–62, 1996.

[111] P. Meikle, J. Hopwood, A. Clague and W. Carey, “Prevalence of lysosomal storage disorders.,” JAMA, vol. 281, no. 3, p. 249–254, 1999.

