## Supplemental Information for "The landscape of functional brain network impairments in late-onset GM2 gangliosidosis"

### S1. Connectivity impairments only within the cerebellum

Because of the specific involvement of the cerebellum in LOGG pathology [1], static functional connectivity (SFC) and variance of dynamic FC (vDFC) impairments only within the cerebellum were additionally assessed as a secondary analysis ($p<0.05$, FDR corrected for ROIs within the cerebellum only). The ROIs were 14 cerebellar regions from the Buckner atlas [2].

SFC impairments only within the cerebellum are reported in **Table S1**. Generally, hypo-connectivity was observed in default mode, somatomotor, and ventral attention networks within the cerebellum. With an uncorrected threshold of $p<0.05$, 62.5% of all intra-cerebellar SFC values were lower in LOGG. This suggests a hyper-connectivity profile (higher SFC) in the cortex and a hypo-connectivity profile (lower SFC) within the cerebellum in LOGG.

**Table S1.** *Static functional connectivity results within the cerebellum only (LOGG vs. control). LOGG group exhibited mainly hypoconnectivity; three of four connections, mainly associated with default mode and somatomotor networks, were reduced in the disease.*

| | Network connectivity | Connection | MNI centroids | p-value | T stat [a] | Effect size |
|---|---|---|---|---|---|---|
| **1** | Somatomotor ↔ Default mode | Cerebellar Culmen R ↔ Cerebellum Crus2 L | (14, -52, -29) ↔ (-24, -73, -37) | 0.0015 | −4.01 | 2.26 |
| **2** | Somatomotor ↔ Default mode | Cerebellar Culmen L ↔ Cerebellum Crus2 L | (-13, -52, -28) ↔ (-24, -73, -37) | 0.0081 | −3.12 | 1.77 |
| **3** | Ventral attention ↔ Default mode | Cerebellar Tonsil L ↔ Cerebellum Crus2 L | (-24, -56, -39) ↔ (-24, -73, -37) | 0.0142 | −2.83 | 1.52 |
| **4** | Ventral attention ↔ Limbic | Cerebellar Tonsil L ↔ Cerebellar Culmen R | (-24, -56, -39) ↔ (17, -49, -36) | 0.0102 | 3.01 | 1.63 |

[a] *Positive T statistic:* Higher connectivity in LOGG compared to controls; *negative T statistic*: lower in LOGG

With the secondary analysis of vDFC impairments only within the cerebellum (**Table S2**), lower variability of connectivity was observed across the board, mainly in cognitive control networks within the cerebellum. With an uncorrected threshold of $p<0.05$, 100% of all intra-cerebellar vDFC values were lower in LOGG. This reconfirms a pattern of lower temporal variability of connectivity both within the cerebellum and partly within the cortex in LOGG.

**Table S2.** *Dynamic functional connectivity results within the cerebellum only (LOGG vs. control). LOGG group exhibited reduced variability of connectivity; all three connections, mainly associated with attention and task control networks, were reduced in the disease.*

| | Network connectivity | Connection | MNI centroids | p-value | T stat [a] | Effect size |
|---|---|---|---|---|---|---|
| **1** | Dorsal attention ↔ Frontoparietal task control | Cerebellum 8 L ↔ Cerebellar Declive L | (-3, -66, -23) ↔ (30, -64, -36) | 0.0032 | −3.61 | 1.93 |
| **2** | Dorsal attention ↔ Visual | Cerebellum 8 L ↔ Cerebellar Vermis L | (-3, -66, -23) ↔ (-17, -63, -46) | 0.0058 | −3.29 | 1.76 |
| **3** | Ventral attention ↔ Limbic | Cerebellar Tonsil L ↔ Cerebellar Culmen L | (-24, -56, -39) ↔ (-16, -49, -35) | 0.0105 | −2.99 | 1.62 |

[a] *Positive T statistic:* Higher variability of connectivity in LOGG vs. controls; *negative T statistic*: lower in LOGG

There also appeared to be slight right-lateralization in the cortical connectivity findings, with 62.5% of the regions involved in impaired SFC or vDFC belonging to the right hemisphere. On the contrary, 86% of all regions involved in intra-cerebellar SFC or vDFC impairments belonged to the left hemisphere. However, with our small sample size, it is possible that these numbers could be driven simply by lateralization of abnormalities in these patients. For example, if 5 of 7 subjects had left-sided ataxia, it might have biased our finding of left cerebellar lateralization and right cerebral lateralization of connectivity abnormalities.

### S2. Associations between fMRI measures and subscales of clinical variables

In addition to the fMRI associations with clinical variables presented in the main text, we assessed the association with subscales of some clinical variables: Friedreich ataxia rating scale (FARS) subscales [3] and cerebellar neuropsychiatric rating scale (CNRS) subscales [4]: attention control – positive and negative symptoms, emotional control – positive and negative symptoms, autism spectrum – positive and negative symptoms, and psychosis spectrum – positive and negative symptoms. It must be noted that the CNRS scale is not clinically validated, and it was obtained merely as a guide to the burden of neuropsychiatric symptoms. Associations with CNRS subscales must thus be interpreted with caution. We however presented it here for the benefit of future research, in case CNRS gets clinically validated in the future.

Since these were tertiary results, multiple comparisons correction was not performed for these associations ($p$<0.05, uncorrected). Additionally, CNRS and its subscales were not considered for the main analysis of fMRI measures vs. clinical variables presented in the main text, and were only chosen for the tertiary analysis presented here.

We found significant associations between certain LOGG imaging measures and some clinical subscales (**Table S3, Figure S1**). Interestingly, the connectivity between ventral and dorsal attention networks was associated with negative attention control, and vDFC with the somatomotor network was associated with ataxia bulbar subscale. Lateral prefrontal (task control) region's aggregate communication with the brain was associated with negative psychosis symptoms. Cognitive control networks featured in all the associations. Figure S1 also shows LOSD patients in a different color, from which it is apparent that these associations were not driven by differences between LOTS and LOSD (we could not demonstrate this statistically because there were only two LOSD patients).

**Table S3.** *Significant associations between subscales of clinical variables and fMRI measures. Negative attention control and psychosis negative symptoms are subscales of CNRS.*

| | Clinical variable | Network | Connection / Node | R-value | $R^2$ value | p-value |
|---|---|---|---|---|---|---|
| | **Static functional connectivity** | | | | | |
| **1** | Negative attention control | Ventral attention ↔ Dorsal attention | TPJ R ↔ Parietal Sup R | −0.80 | 0.63 | 0.032 |
| | **Variance of dynamic functional connectivity** | | | | | |
| **2** | FARS (bulbar subscale) | Somatomotor ↔ Salience | Postcentral L ↔ Suppl. Motor Area L | −0.80 | 0.65 | 0.029 |
| | **Complex network measures** | | | | | |
| **3** | Psychosis negative symptoms | *Local efficiency*: Frontoparietal task control | Frontal Mid L | −0.84 | 0.71 | 0.018 |

FARS = Friedreich ataxia rating scale, TPJ = temporo-parietal junction, sup = superior, mid = middle, suppl = supplementary.

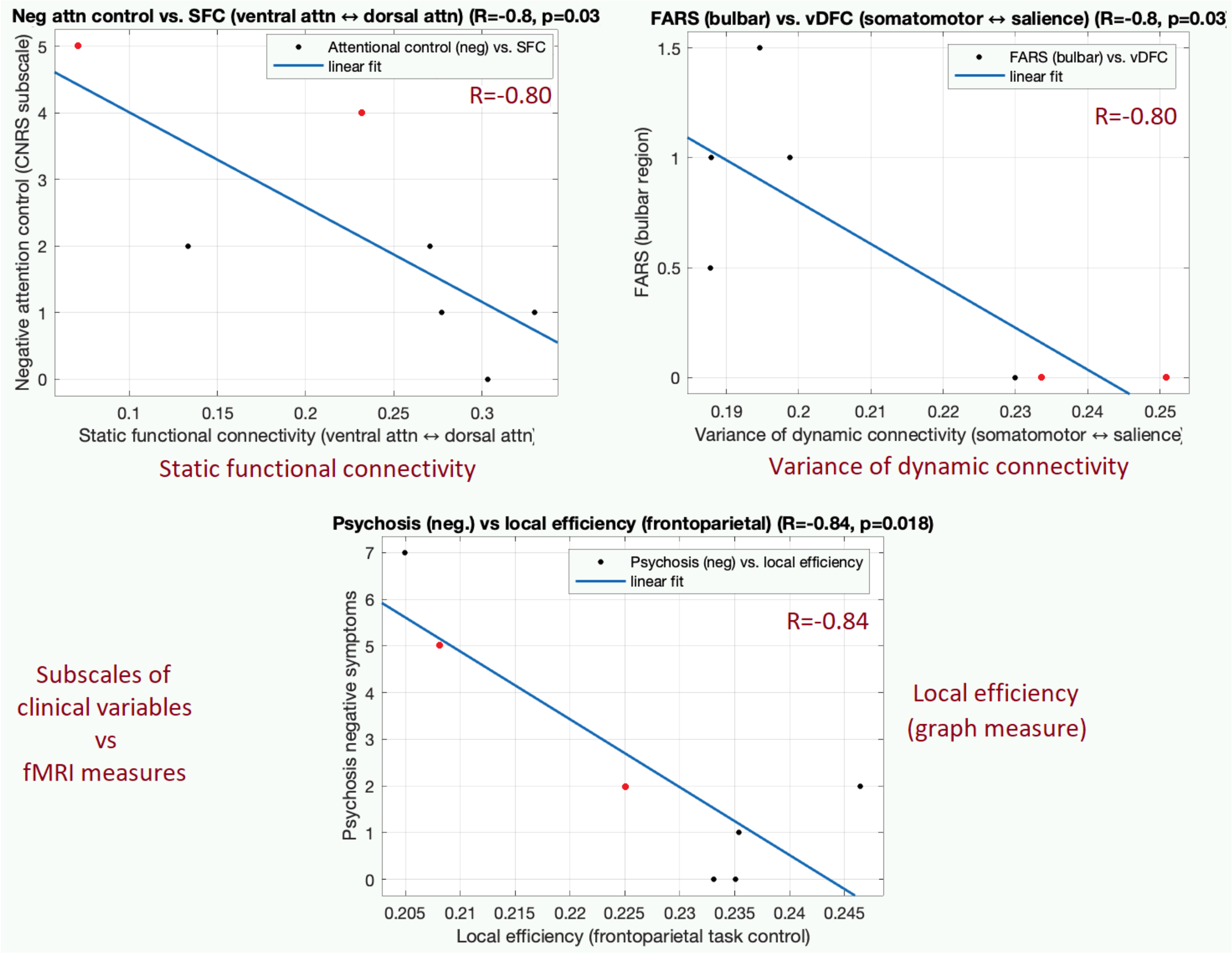


**Figure S1**. *Associations between fMRI measures and subscales of clinical variables. Top-left: SFC vs. negative attentional control (CNRS subscale) (R=−0.80, p=0.032). Top-right: vDFC vs. FARS bulbar subscale (R=−0.80, p=0.029). Bottom: local efficiency vs. psychosis negative symptoms (CNRS subscale) (R=−0.84, p=0.018).* The two LOSD patients are shown as red points. *FARS = Friedreich ataxia rating scale.*

### Supplemental References

[1] O. E. Rowe, D. Rangaprakash, A. Weerasekera, N. Godbole, E. Haxton, P. F. James, C. D. Stephen, R. L. Barry, F. S. Eichler and E. M. Ratai, "Magnetic resonance imaging and spectroscopy in late-onset GM2-gangliosidosis.," *Molecular genetics and metabolism,* vol. 133, no. 4, p. 386–396, 2021.

[2] R. Buckner, F. Krienen, A. Castellanos, J. Diaz and B. Yeo, "The organization of the human cerebellum estimated by intrinsic functional connectivity," *J Neurophysiol.,* vol. 106, no. 5, pp. 2322-45, 2011.

[3] S. Subramony, W. May, D. Lynch, C. Gomez, K. Fischbeck, M. Hallett, P. Taylor, R. Wilson, T. Ashizawa and C. A. Group, "Measuring Friedreich ataxia: Interrater reliability of a neurologic rating scale.," *Neurology,* vol. 64, no. 7, p. 1261–1262, 2005.

[4] F. Hoche, X. Guell, J. Sherman, M. Vangel and J. Schmahmann, "Cerebellar Contribution to Social Cognition.," *Cerebellum,* vol. 15, no. 6, p. 732–743, 2016.
